# Interoperability of phenome-wide multimorbidity patterns: a comparative study of two large-scale EHR systems

**DOI:** 10.1101/2024.03.28.24305045

**Authors:** Nick Strayer, Tess Vessels, Karmel Choi, Siwei Zhang, Yajing Li, Lide Han, Brian Sharber, Ryan S Hsi, Cosmin A Bejan, Alexander G. Bick, Justin M Balko, Douglas B Johnson, Lee E Wheless, Quinn S Wells, Elizabeth J Philips, Jill M Pulley, Wesley H Self, Qingxia Chen, Tina Hartert, Consuelo H Wilkins, Michael R Savona, Yu Shyr, Dan M Roden, Jordan W Smoller, Douglas M Ruderfer, Yaomin Xu

## Abstract

**Background:** Electronic health records (EHR) are increasingly used for studying multimorbidities. However, concerns about accuracy, completeness, and EHRs being primarily designed for billing and administrative purposes raise questions about the consistency and reproducibility of EHR-based multimorbidity research.

**Methods:** Utilizing phecodes to represent the disease phenome, we analyzed pairwise comorbidity strengths using a dual logistic regression approach and constructed multimorbidity as an undirected weighted graph. We assessed the consistency of the multimorbidity networks within and between two major EHR systems at local (nodes and edges), meso (neighboring patterns), and global (network statistics) scales. We present case studies to identify disease clusters and uncover clinically interpretable disease relationships. We provide an interactive web tool and a knowledge base combining data from multiple sources for online multimorbidity analysis.

**Findings:** Analyzing data from 500,000 patients across Vanderbilt University Medical Center and Mass General Brigham health systems, we observed a strong correlation in disease frequencies ( Kendall’s *τ* = 0.643) and comorbidity strengths (Pearson *ρ* = 0.79). Consistent network statistics across EHRs suggest similar structures of multimorbidity networks at various scales. Comorbidity strengths and similarities of multimorbidity connection patterns align with the disease genetic correlations. Graph-theoretic analyses revealed a consistent core-periphery structure, implying efficient network clustering through threshold graph construction. Using hydronephrosis as a case study, we demonstrated the network’s ability to uncover clinically relevant disease relationships and provide novel insights.

**Interpretation:** Our findings demonstrate the robustness of large-scale EHR data for studying phenome-wide multimorbidities. The alignment of multimorbidity patterns with genetic data suggests the potential utility for uncovering shared biology of diseases. The consistent core-periphery structure offers analytical insights to discover complex disease interactions. This work also sets the stage for advanced disease modeling, with implications for precision medicine.

**Funding:** VUMC Biostatistics Development Award, the National Institutes of Health, and the VA CSRD

## Introduction

Multimorbidity, the coexistence of multiple health conditions within an individual, poses significant challenges for personalized care and hinders patient-centric medicine.^1,2^ Electronic health records (EHRs) offer a rich data resource for investigating phenome-wide multimorbidity patterns at scale^3–5^, capturing real-world disease interactions. Understanding multimorbidity patterns is essential for unraveling disease heterogeneity, uncovering shared etiology, enhancing precision in disease risk assessments, and optimizing decision support tailored to individual needs ^6–9^.

Network analysis has emerged as a pivotal tool for large-scale multimorbidity research^10^. By modeling diseases as nodes and their co-occurrences as edges, this method provides an intuitive and efficient approach to explore intricate disease relationships. However, concerns remain about the reproducibility of such analyses, stemming from limitations in the accuracy and completeness of EHR data and its primary design for billing and administrative purposes.

In this study, we address these concerns through a comparative analysis of multimorbidity networks derived from two large-scale EHR systems. We assessed the consistency of multimorbidity patterns both within and across the systems, employing graph-theoretical analyses to derive novel insights and enhance knowledge discovery. Our findings demonstrate the remarkable robustness of multimorbidity network analyses across EHRs at multiple scales, from local-scale disease frequencies and comorbidity strengths to meso-scale core-periphery and clustering structures, and up to global network statistics (Figure 1A). These networks effectively identify clinically relevant disease clusters, and a case study on hydronephrosis revealed their capability to uncover both known and potentially novel causative disease associations. Additionally, we have developed an interactive web-based tool to facilitate the exploration of multimorbidity patterns and their comparisons across systems. Our analyses indicate the strong interoperability of network analysis leveraging extensive diagnostic information from EHRs and affirm the reliability of our multimorbidity network modeling strategy.

**Figure 1.**
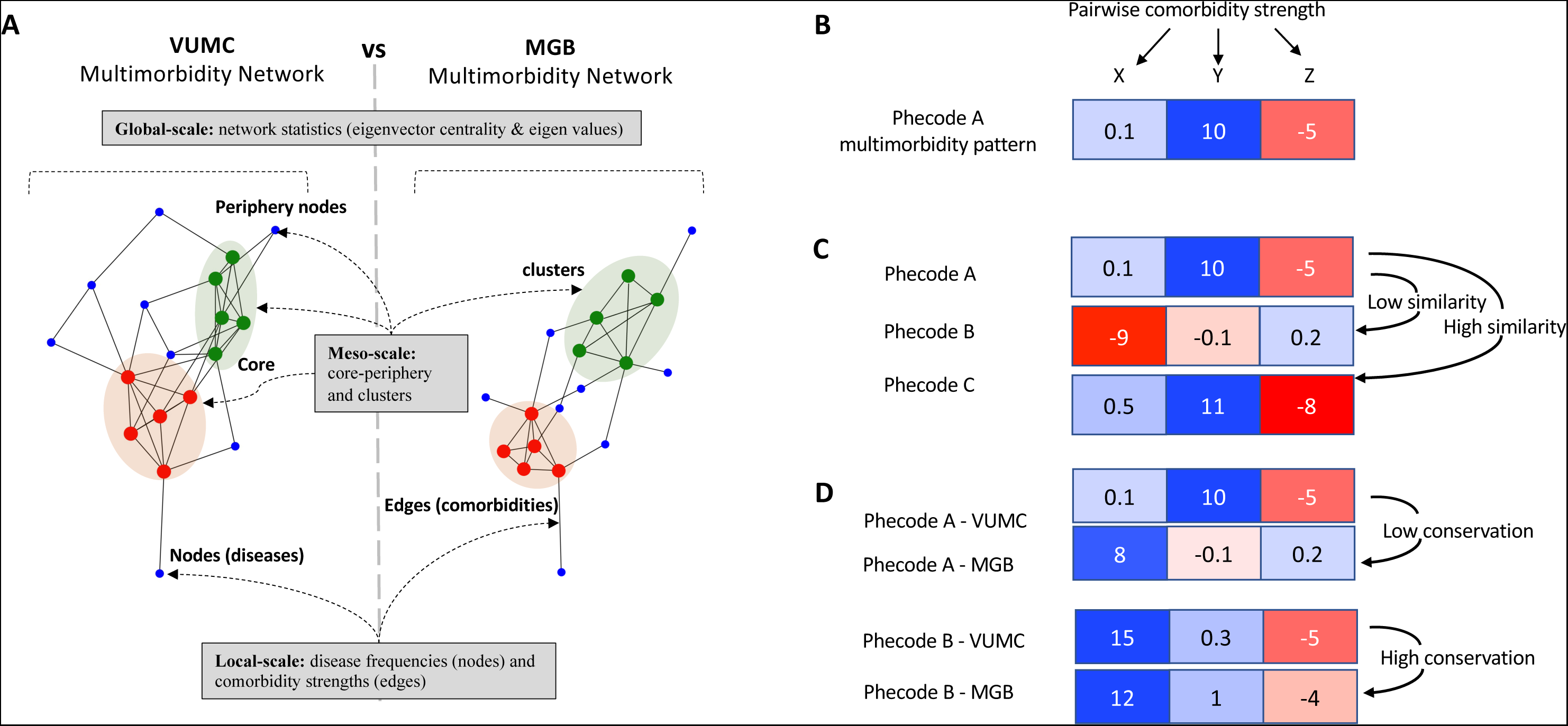
Schematic Demonstration of Multimorbidity Networks, Connection Patterns, Similarity, and Conservation. **A.** Multimorbidity networks represent diseases (nodes) and their connections (edges), where edges represent pairwise comorbidity strengths. Analysis of multimorbidity networks can occur at the global scale where network statistics are utilized to characterize the overall shape and connectivity of the entire network, at the meso-scale where it describes intermediate structures such as core-periphery patterns (where diseases cluster in a densely connected core or a less connected periphery) or other distinct disease cluster patterns, and at the local scale where the focus is on the properties of individual diseases (nodes) and their connections (edges). **B.** A single disease’s multimorbidity pattern (e.g., Phecode A) is defined by its comorbidity strengths with other diseases (Phecode X, Y, Z). **C.** Multimorbidity similarity assesses how closely two diseases’ comorbidity patterns align. Phecode A is more similar to C than B. **D.** Conservation examines the consistency of multimorbidity patterns across systems. Phecode B’s pattern is highly conserved between VUMC and MGB, while Phecode A’s shows less conservation.

## Methods

### EHR and disease phenome

We conducted a prevalence study to characterize phenome-wide disease comorbidity patterns across two large-scale electronic health record (EHR) systems. Individual-level diagnostic code data were extracted for 250,000 randomly selected patients from each of Vanderbilt University Medical Center (VUMC) and Mass General Brigham (MGB)’s EHR systems, which comprised 2.2 million and 1.8 million patients at the time of data extraction, respectively. The selection of 250,000 patients from each institution was intended to balance a representative sample with computational feasibility and costs. The sampled patients’ longitudinal records were then collapsed to the number of occurrences of ICD9 and ICD10 codes. These code counts were mapped to phecodes v1.2 using the PheWAS R package^11^. Demographic data, including patient age at sample extraction date, EHR age (patient age at last recorded visit), sex, self-identified race, and the logarithm of the disease burden (number of unique phecodes), were extracted for model adjustment. These covariates help account for age-related disease patterns, healthcare utilization differences (indicated by EHR age and burden), and potential biases related to demographic differences reflected in sex and race. The covariate adjustments address the fact that the observed multimorbidity patterns are driven by demographic factors or variations in healthcare utilization^1,12–14^. The study was approved by the Vanderbilt University Medical Center (VUMC) Institutional Review Board (IRB# 172041) and by the Mass General Brigham (MGB) Institutional Review Board (IRB# 2018P002642).

### Dual regression analysis of disease comorbidities

We applied a dual logistic regression analysis strategy to characterize pairwise comorbidity strengths for each phecode pair. Previous studies demonstrated the utility of regression models for large-scale comorbidity analysis^15^. This approach allows us to adjust for confounding factors that vary across institutions or populations, as well as differences in disease prevalence that could influence the accuracy of observed comorbidity frequencies. Our dual regression analysis includes two regression models for each pair of phecodes (e.g., phecode A and B):

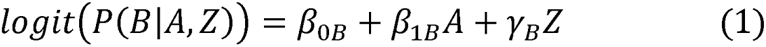

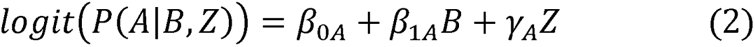

where *Z* represents covariates including patient age at the event, hospital usage in years, sex, self-identified race and the logarithm of disease burden. We extracted and averaged the test statistics (Wald statistics) of the *β*_1A_ and *β*_1B_ coefficients from the two regressions to generate a symmetric score for each phecode pair to normalize the phecode effects which we call pairwise comorbidity strength (PCS) of the phecode pair, or simply comorbidity strengths of the phecode pair.

This approach simplifies the complex nature of multimorbidity, enabling a structured evaluation of both disease association strengths and statistical significances while adjusting for potential confounders. This ensures robust findings, distinguishing true associations from spurious correlations within large EHR datasets and promoting generalizability across diverse populations. The detailed methodology for the regression analyses is provided in the Supplementary Materials (see Supplementary A1)

### Network model of phenome-wide multimorbidities

We model phenome-wide disease multimorbidities as undirected, weighted networks where diseases are nodes, and edges represent the pairwise comorbidity strengths derived from our dual regression analysis. Separate networks were constructed for the VUMC and MGB EHR systems to enable comparative analysis and assess the consistency of multimorbidity patterns. This network modeling approach provides an intuitive foundation for understanding intricate multi-disease interactions, facilitates analyses such as disease cluster identification and disease progression modeling, and quantifies disease importance within the multimorbidity network using graph-theoretic principles. Our approach is computationally efficient, easy to visualize, and serves as a strategic starting point for characterizing complex disease-disease relationships. It balances simplicity with the ability to derive essential insights about complex disease relationships.

### Structural equivalence in multimorbidity networks

We define a phecode’s "multimorbidity pattern" as its profile of comorbidity strengths to all other phecodes within the network (Figure 1B). To quantify the similarity between phecodes, considering both the magnitude of comorbidity strength and pattern of connections with neighboring nodes, we use a correlation-based structural equivalence measure^16^. Two phecodes are structurally equivalent if their multimorbidity patterns exhibit a high correlation, indicating similar relationships with other diseases within the network (Figure 1C, Supplementary A2). This approach characterizes nuanced meso-scale structures (clustering pattern) and positional roles of nodes within their neighboring nodes. It is often hypothesized that structurally equivalent nodes in a network will be similar in other ways such as sharing common mechanisms or functions.

Since our multimorbidity networks are constructed using identical phecode mappings, we can directly compare the structure equivalence measurements across two EHR systems. Therefore, we define the "conservation " of each phecode as the correlation between its comorbidity strength vectors in the VUMC and MGB networks (Figure 1D, Supplementary A2). High conservation scores indicate consistent multimorbidity patterns across EHR systems, suggesting that the underlying mechanisms driving disease-disease relationships are transferable across the systems.

This intra- and inter-system structural equivalence analysis offers a novel approach to investigate multimorbidity relationships. By focusing on connection patterns with others, it transcends simple pairwise comorbidities to uncover complex disease interactions, while facilitating the evaluation of both the generalizability of findings and the transferability of implied analysis strategies across EHR systems. The detailed methodology of structural equivalence analysis and its application to analyzing multimorbidity patterns within and between systems is provided in the Supplementary Materials (see Supplementary A2).

### Spectral analysis of multimorbidity network topology

We use network centrality measures to assess the global-scale topological consistency of multimorbidity networks across institutions. Eigenvector centrality, calculated from the leading eigenvector of the network adjacency matrix, quantifies a node’s importance based on the number, strength, and importance of its connections in the network. Analyzing and comparing the distribution of centrality measures across the networks reveals the overall structure and their consistency^17^. Specifically, we consider the multimorbidity network as an undirected network of phecode nodes (phecode *v*1.2). The eigenvector centrality *x_i_* of disease *i* is the *i*^th^ element of the leading eigenvetor ***x*** of the network adjacency matrix A, such that,

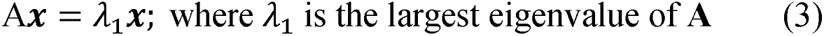

To evaluate meso-scale structure of the multimorbidity networks, we used eigengap heuristic^18^. This metric, defined as 1−*λ*_2_/*λ*_1,_ where *λ*_1_ and *λ*_2_ are the largest and second largest eigenvalues of the network adjacency matrix A, respectively, assesses how closely a network resembles an ideal core-periphery model^19^ (like a threshold graph). Higher eigengap values indicate a densely connected core, suggesting a prominent core-periphery structure. For a more detailed discussion of spectral analysis techniques and applications to multimorbidity networks, please see Supplementary Materials A3.

### Construction of consensus multimorbidity network

To facilitate cross-institutional analysis, we constructed a consensus multimorbidity network. Pairwise comorbidity strengths in this consensus network were calculated as weighted averages of the corresponding comorbidity strengths from the VUMC and MGB networks. Weights were proportional to the number of shared patients exhibiting each comorbidity pair in the respective system. For phecodes A and B, the combined pairwise comorbidity strength score (PCS) is calculated as follows:

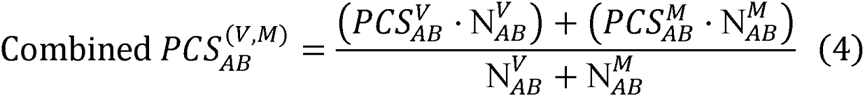

where 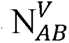 and 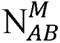 are number of shared patients between phecodes A and B in VUMC (V) and MGB (M) cohorts, respectively. This weighting scheme prioritizes comorbidity estimates supported by larger patient populations, increasing the reliability of the consensus network.

### Interactive online exploration of phenome-wide multimorbidities

We developed an interactive web application (https://prod.tbilab.org/PheMIME/) to facilitate the exploration of multimorbidity patterns across multiple EHR systems ^20^. This PheMIME tool allows users to visualize pairwise comorbidity patterns across institutions, examine the consistency of multimorbidity relationships across diverse EHR systems, and explore subgraph structures using associationSubgraphs ^21^. This application provides researchers an intuitive interface to explore complex multimorbidity data, facilitating insights into disease relationships and the reproducibility of findings across different healthcare datasets. For a detailed explanation of the associationSubgraphs algorithm and its connection to the theoretical properties of threshold graphs, please refer to Supplementary Materials A4 and A5.

## Results

### High concordance of disease frequencies and comorbidity strengths across EHR systems

We extracted demographic and clinical data (phecodes) for 250,000 randomly selected patients from de-identified EHRs at two healthcare systems (see Methods). The patient populations differed slightly in demographics, with VUMC having a lower proportion of females (55%) compared to MGB (58%), and a higher proportion of white race (81%) compared to MGB (74%). Additionally, VUMC exhibited a younger median age (42.7 years) compared to MGB (52.6 years) likely due to a larger pediatric population (Table 1). Despite these demographic variations, disease frequencies exhibited a strong positive correlation across the phenome (Kendall’s τ=0.643, p < 2.2e-16). We observed higher diagnostic frequencies at VUMC, particularly within the "Sense Organs" category (Figure S4).

**Table 1.**
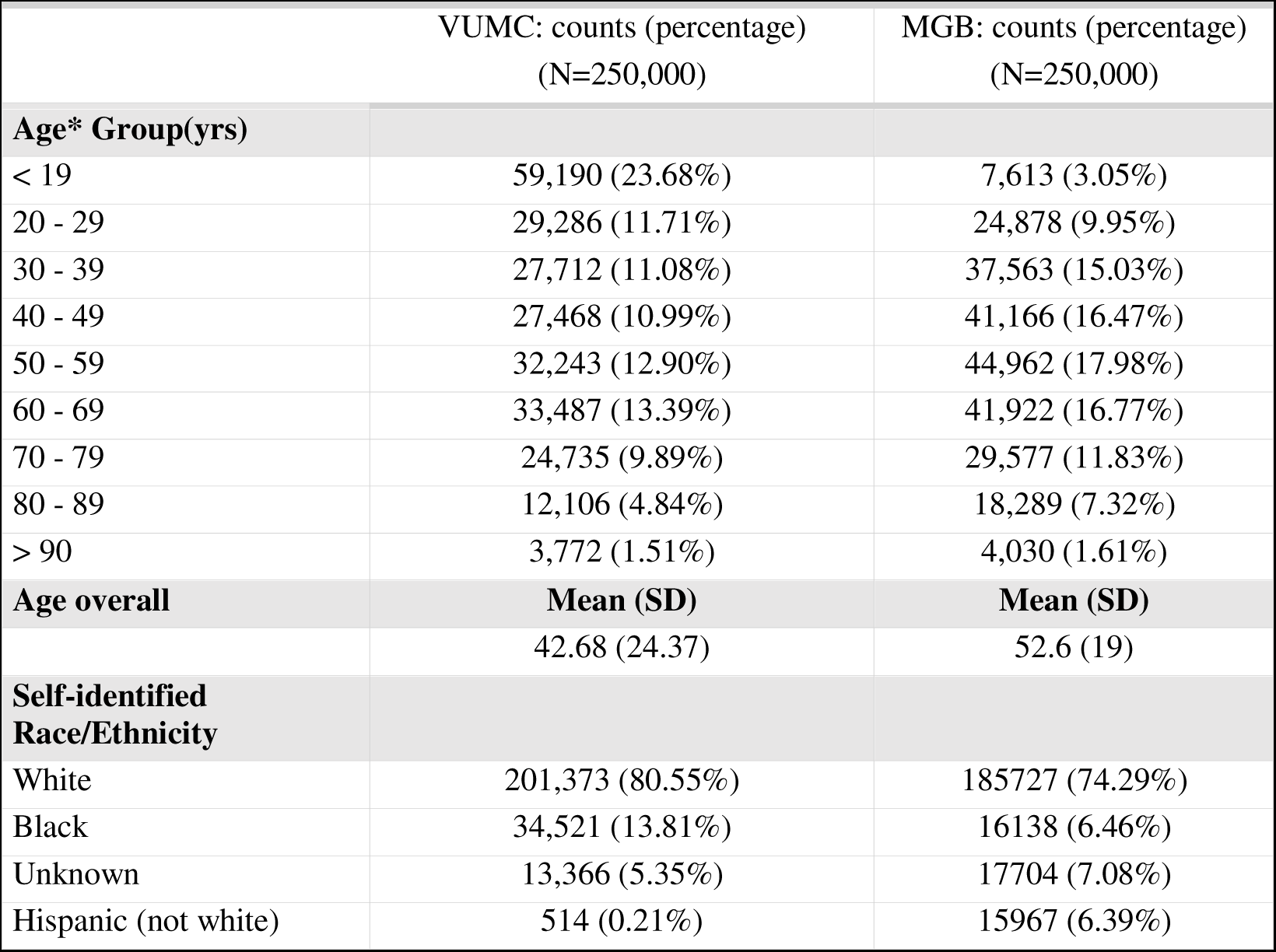
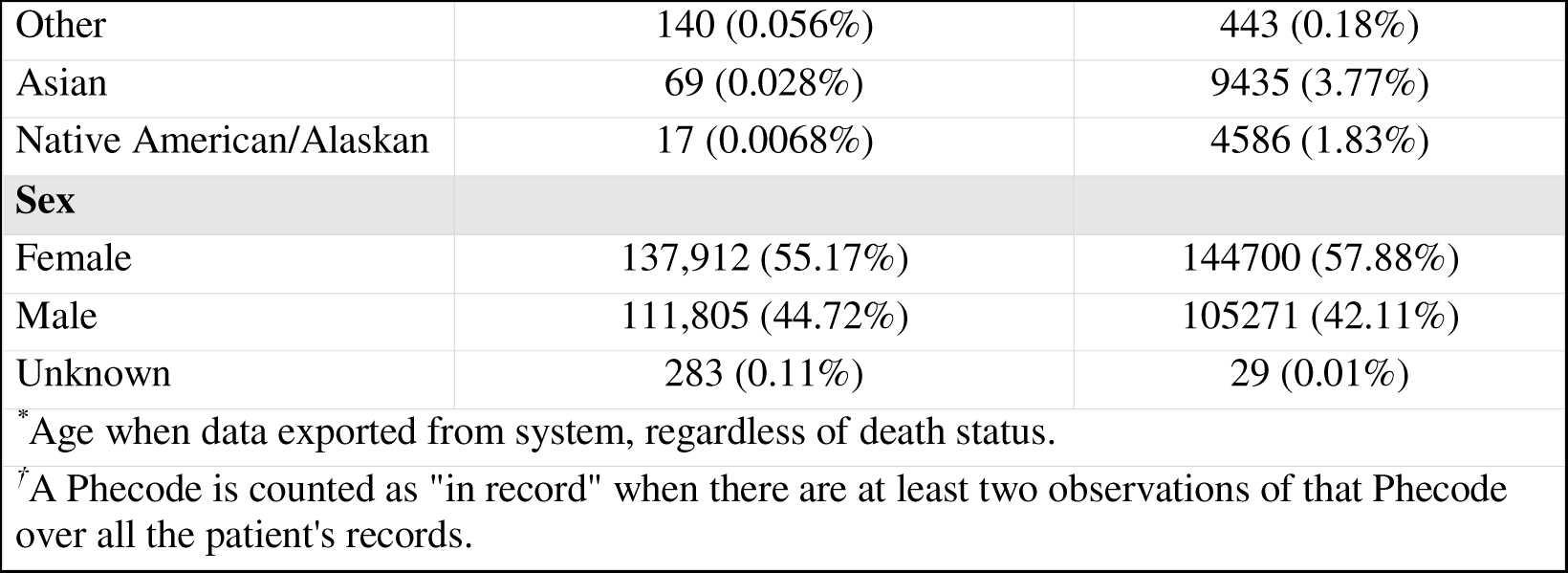
Demographics of EHR patient cohorts.

We then calculated pairwise comorbidity strengths for all phecode pairs using the dual logistic regression approach (see Methods). The comorbidity strength scores demonstrated high concordance between the two EHRs (Pearson ρ=0.79; 95% CI: 0.787 – 0.792, Figure 2A). This indicates that the patterns of disease co-occurrence are remarkably consistent after adjusting for variations in patient populations and potential differences in healthcare practices.

**Figure 2.**
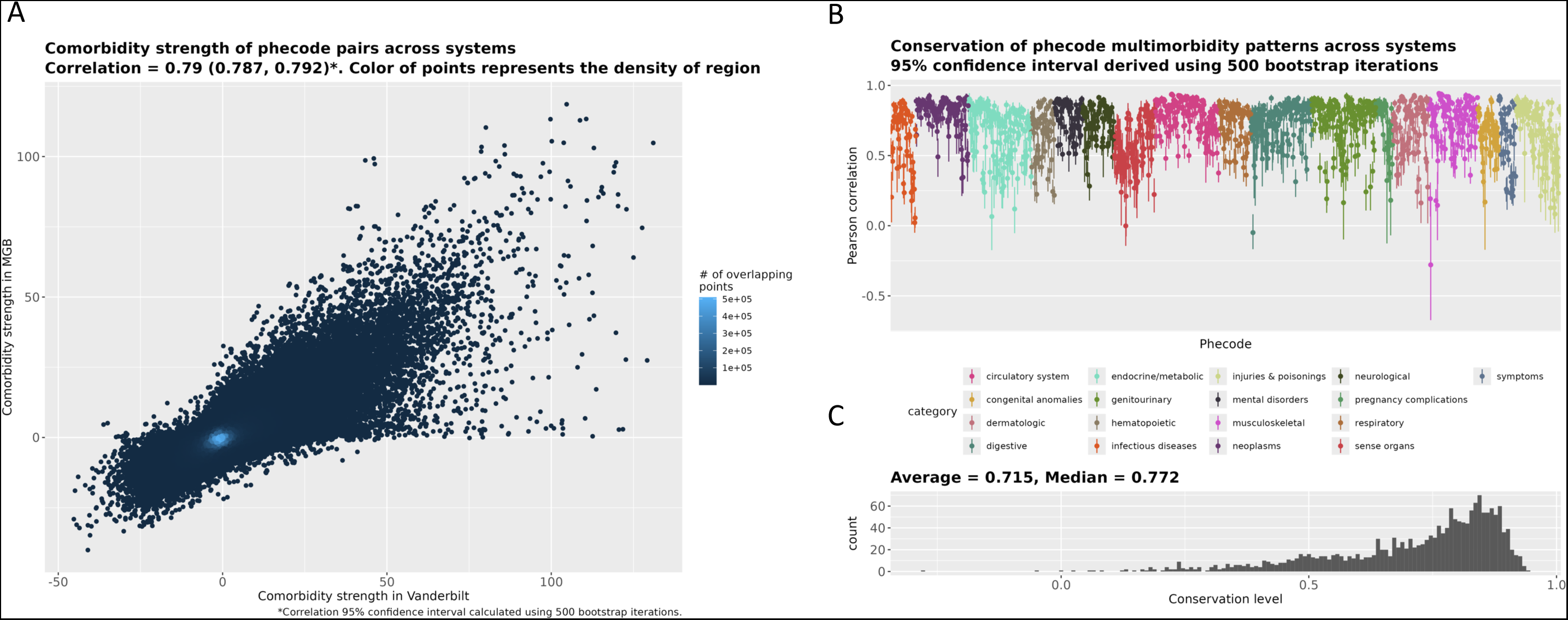
High Conservation of Multimorbidity Patterns Across EHR Systems. **A.** Scatterplot demonstrates a strong correlation between pairwise comorbidity strengths for all common phecode pairs in VUMC and MGB systems. This indicates that diseases exhibiting high (or low) comorbidity in one EHR tend to show a similar pattern in the other. **B.** Conservation of phecodes’ multimorbidity patterns, measured by Pearson correlation coefficient-based structural equivalence, assesses the consistency if a disease’s connections with others within the multimorbidity network aligns across EHR systems. Dots represent bootstrap means, with confidence intervals reflecting uncertainty. High conservation for most phecodes demonstrates that their connectivity patterns within the network are largely consistent. **C.** The distribution of bootstrap means skews towards 1, highlighting strong phecode conservation across EHR systems.

### Strong conservation of multimorbidity patterns across EHRs, with variations offering insights

We assessed the conservation of multimorbidity patterns across the two EHR systems using correlation-based structural equivalence (see Methods, Supplementary A2). High correlation scores indicate that a disease consistently co-occurs with the same set of diseases across EHRs. Specifically, we calculated Pearson correlations of phenome-wide multimorbidity patterns for each phecode with 500 bootstrap iterations to assess variability (Figure 2B). Overall, we observed strong conservation (median correlation = 0.772, Figure 2C). "Spinal stenosis" had the most conserved patterns (correlation = 0.942, CI = 0.921-0.958), while "Dental abrasion, erosion, and attrition" was the least conserved (correlation = -0.051, CI = -0.182-0.078). Examining conservation across categories, we found "Sense Organs" phecodes exhibited the lowest average conservation, while "Neoplasms" were the most conserved (Figure S5). These variations in conservation likely reflect differences in site-specific healthcare practices and patient populations.

### Consistent global structure of multimorbidity networks across EHRs

We assessed the global-scale structural consistency of multimorbidity networks across institutions using leading eigenvector centrality (see Methods, Supplementary A3), a metric that quantifies a disease node’s importance based on the number, strength, and centrality of its connections. Our analysis revealed a remarkably high correlation in eigenvector centralities between VUMC and MGB (Spearman correlation = 0.902; 95% CI: 0.889 – 0.914, Figure 3A), indicating a consistent global network structure across systems.

**Figure 3.**
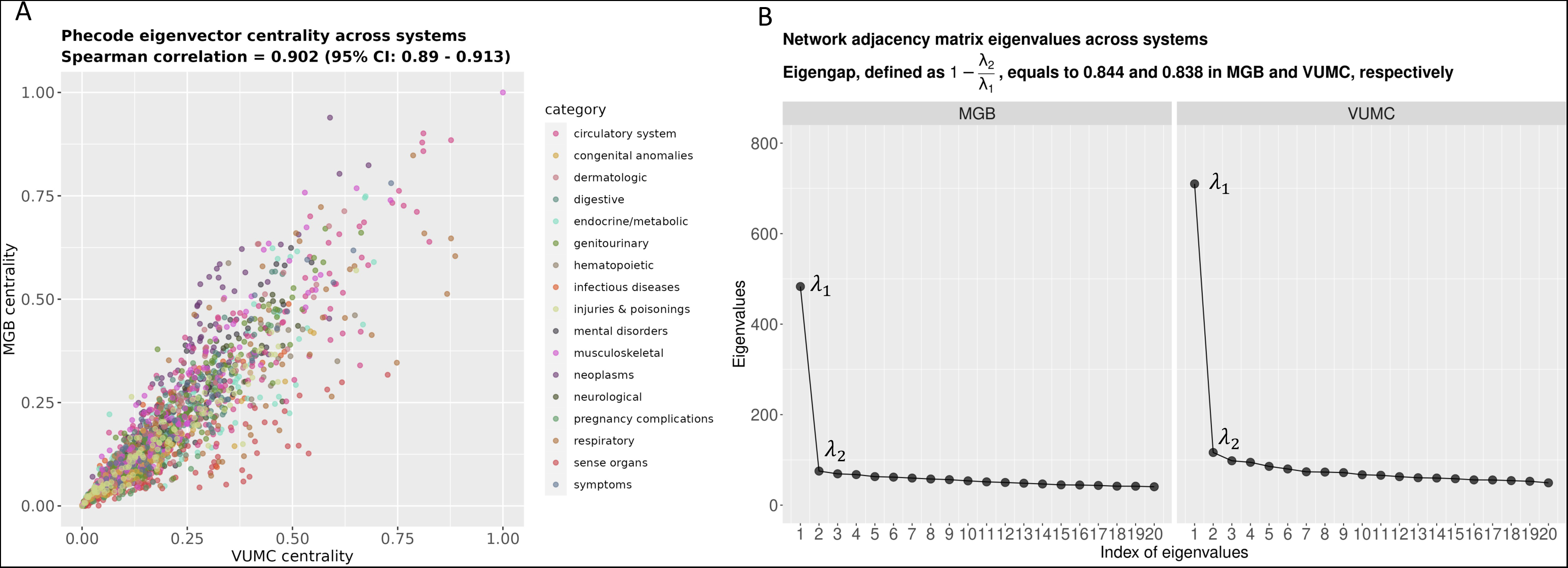
Consistent Spectral Properties of Multimorbidity Networks. **A.** Eigenvector centrality scores, which reflect a disease’s importance within the network, exhibit a strong correlation across the two EHR systems. This indicates that most diseases maintain similar topological positions within their respective multimorbidity networks. Notable differences exist in the "sense organs" category, likely reflecting dataset-specific factors. **B.** Similar eigengap values across systems suggest a consistent core-periphery structure, supporting the robust conservation of meso-scale structures of multimorbidity networks.

Despite this overall consistency, we observed nuanced differences in centrality for specific disease categories. "Sense Organ" (diseases and disorders of the eyes and ears) and "Respiratory" phecodes (e.g., "Cataract" and "Acute Sinusitis") tended to be more central in the VUMC network, while "Neoplasms" were more central in the MGB network (Table S3, Figure S7). These patterns are further explored in Supplementary A9, where we analyze eigen-centrality distributional differences across categories. Notably, only "Neoplasms," "Musculoskeletal," and "Dermatologic" phecodes were, on average, more central in MGB.

This high consistency of centrality measures across VUMC and MGB networks, despite variations in patient populations and healthcare practices, suggests a high degree of robustness in the overall structure of multimorbidity networks, and strengthens the generalizability of network-based insights into multimorbidity patterns using large-scale EHR data.

### Spectral analysis reveals core-periphery structure of multimorbidity networks

Spectral analysis revealed prominent core-periphery structures within the multimorbidity networks for both VUMC and MGB. This is evidenced by their large eigengaps (0.836 and 0.844) and consistently high eigenvector centralities across systems (see Methods, Figure 3A&B). In a core-periphery network, a densely connected core exists where diseases exhibit strong interconnections, while a sparsely connected periphery contains diseases with weaker links. Our UMAP representation (Figure 4) visually supports this interpretation, with musculoskeletal, metabolic, circulatory, and injuries & poisoning diseases enriched in the core.

**Figure 4.**
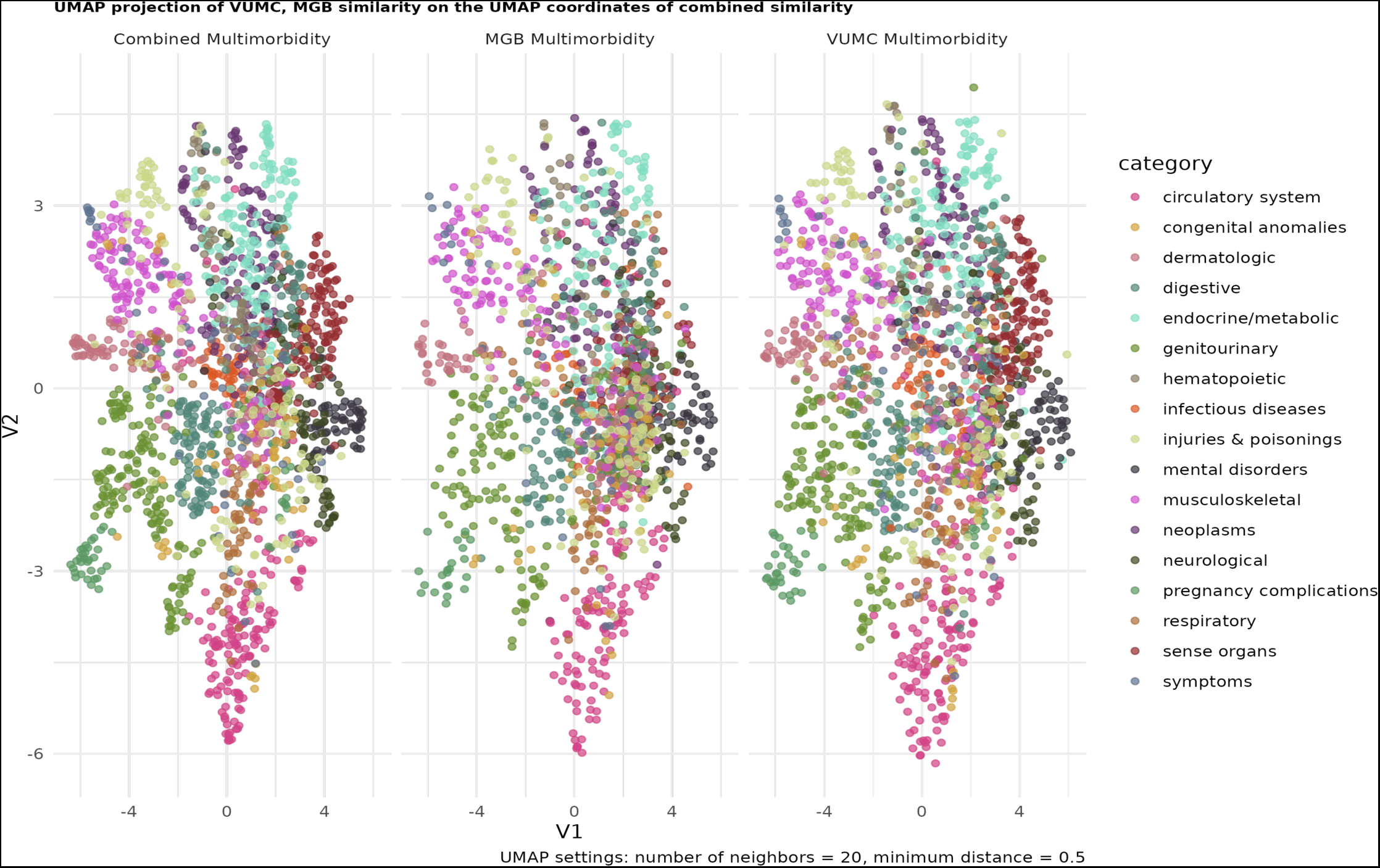
UMAP Visualization Reveals Consistent Multimorbidity Network Structure with Dataset-specific Variation. UMAP projection of multimorbidity networks reveals remarkable consistency in their overall structure across EHR systems. Both networks exhibit a prominent core-periphery structure, and phecodes cluster by their disease categories (e.g., circulatory, musculoskeletal), with consistent sub-clustering patterns within these categories. Despite overall consistency, dataset-specific differences exist. For example, the "sense organs" cluster that is more prominent in VUMC, likely reflecting factors specific to that dataset.

This core-periphery structure implies a strategic analytical approach: isolating central disease clusters can identify more commonly occurring diseases in the population with shared mechanisms and functional overlaps, while investigating peripheral diseases could uncover more specific etiologies present only in subsets of patients. Furthermore, the networks’ proximity to threshold graphs enables efficient search algorithms: applying the associationSubgraphs algorithm allowed us to dynamically cluster the multimorbidity networks, identifying subgraphs (disease clusters) even in cases where not all diseases within the cluster exhibit uniformly strong pairwise comorbidities, potentially uncovering broader and more clinically relevant disease clusters (see Supplementary A4 and A5).

### Multimorbidity patterns and genetic correlations of diseases strongly align

Genetically correlated diseases often share common pathophysiological mechanisms, potentially leading to elevated comorbidities. This suggests a link between multimorbidity patterns and shared disease etiology ^22,23^. To investigate this connection, we analyzed the genetic correlation of 15 common, heritable phenotypes (Supplementary A13 & A14). Among the 105 phenotype pairs, 28 (26.7%) showed substantial genetic correlations (>0.5), including well-established pairs like "Coronary Atherosclerosis" and "Myocardial Infarction." We compared pairwise comorbidity strengths and multimorbidity similarity measures with their corresponding genetic correlations. Results revealed significant positive associations (Pearson correlations of 0.59 and 0.62, respectively; Figure 5). Notably, multimorbidity similarity demonstrated a stronger linear relationship with genetic correlation, suggesting it may be a more sensitive indicator of potential shared disease etiology, especially when direct comorbidity is weak.

**Figure 5.**
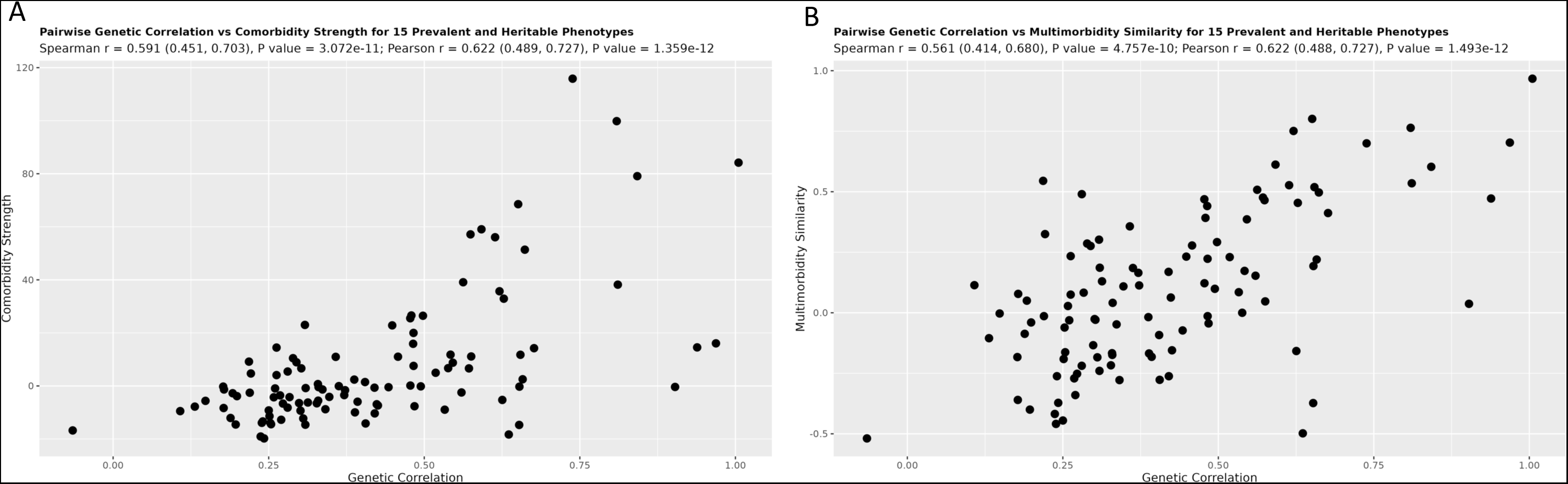
Comorbidity Strength and Multimorbidity Similarity Aligns with Genetic Correlation. Scatterplots demonstrate significant positive associations between phenotypic similarity (based on multimorbidity patterns) and genetic correlations across 15 prevalent, heritable phenotypes. **A.** Pairwise comorbidity strength exhibits a significant positive correlation with genetic correlation. This suggests that diseases with higher comorbidity tend to share more genetic risk factors. **B.** Multimorbidity similarity shows a strong positive association with genetic correlation. This indicates that multimorbidity similarity might be a sensitive indicator of shared underlying disease mechanisms, even in cases where direct comorbidity is not as pronounced.

### Multimorbidity subgraphs identify robust disease clusters

We applied the associationSubgraphs method ^21^ to our multimorbidity networks to identify disease clusters, or "subgraphs", representing groups of diseases with high co-occurrence rates. The consistent structural equivalence observed across different multimorbidity networks (Figure 2, Supplementary Figure S5) provides a strong theoretical basis for identifying clusters that are likely to be robust across different EHR systems and populations (see Methods and Supplementary A5). We identified several prominent disease condition clusters (Figure 6, Supplementary Tables S5-S7) that align with and refine previously reported condition clusters^24^. Examples include the cardiometabolic cluster (encompassing cardiovascular, metabolic diseases, and others), the mental health cluster (including mood disorders, anxiety, bipolar disorder, schizophrenia, substance use disorders, and others), and the musculoskeletal cluster (featuring conditions like back pain, fractures, sprains, and various joint disorders, and others). Additionally, we identified several large clusters (15+ phecodes) related to cancer, dermatological, reproductive, neurological, and eye disorders. Prior research links some of these clusters, like the mental health cluster, with poor outcomes and increased healthcare costs ^25^.

**Figure 6.**
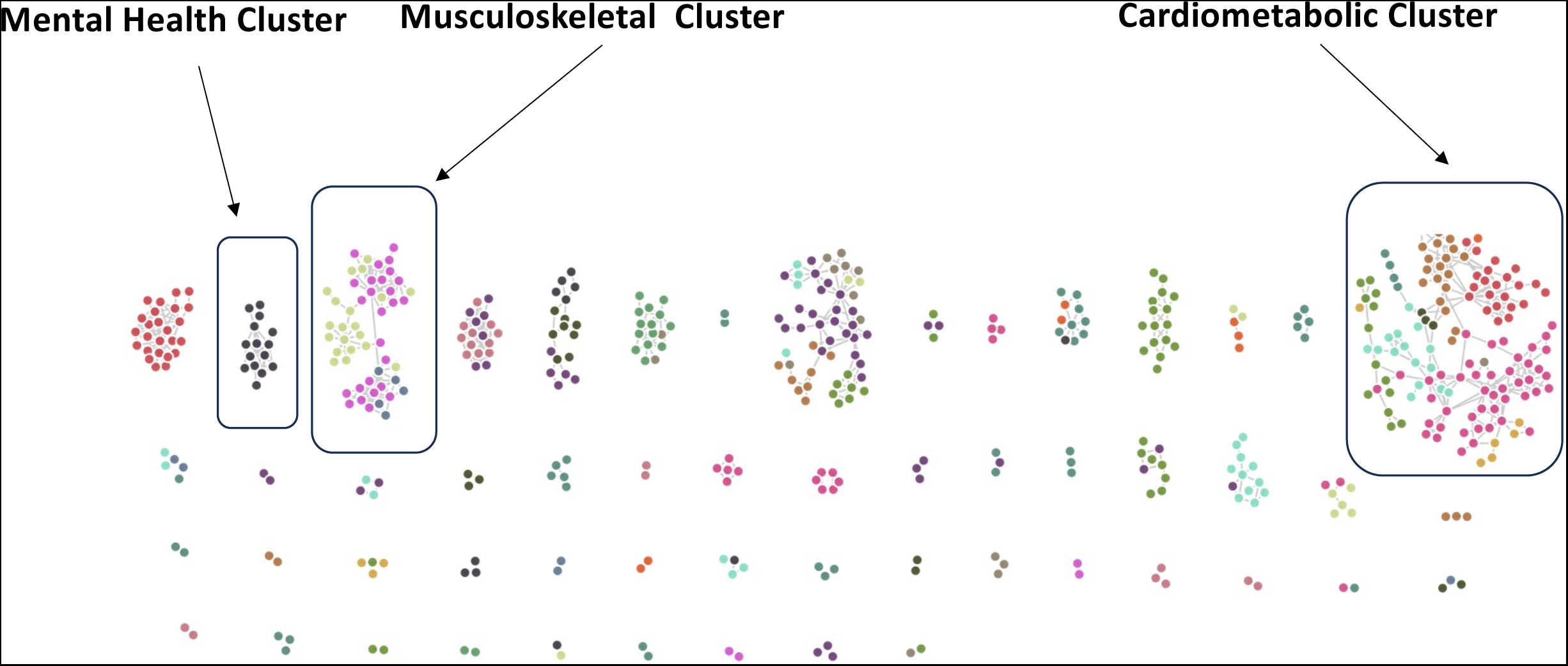
Multimorbidity Network Analysis Identifies Robust Disease Clusters. AssociationSubgraphs analysis of the consensus phenome-wide multimorbidity network reveals several prominent disease clusters (or "condition clusters") that align with those consistently found across multiple studies. Highlighted clusters include the Cardiometabolic Cluster (encompassing cardiovascular and metabolic diseases), the Mental Health Cluster (including mood disorders, depression, anxiety, and other mental health conditions), and the Musculoskeletal Cluster (featuring musculoskeletal disorders and injuries).

### Multimorbidity networks uncover disease relationships: A Hydronephrosis case study

We used hydronephrosis as a case study to demonstrate the utility of multimorbidity networks in revealing known and potentially novel disease relationships. Hydronephrosis is a kidney condition resulting from urinary tract obstruction ^26^. We observed a strong conservation of multimorbidity patterns (conservation value: 0.852; 95% CI: 0.792 - 0.895, Figure 7A) and identified the strongest and most consistent comorbidities associated with hydronephrosis, including established causes like obstructing stones in the ureter, vesicoureteral reflux, congenital defects^27^ (Figure 7A & B, Tables S8 & S9). Additionally, we identified uncommon causes of hydronephrosis, such as cancers, urethral stricture, and abnormal renal vasculature ^28–30^. Interestingly, the analysis also highlighted several mineral metabolism disorders associated with hydronephrosis, although the exact mechanisms linking these conditions remain unclear. These findings suggest the network’s potential to identify novel disease associations for further investigation. These results can be explored interactively using our online application (https://prod.tbilab.org/PheMIME/).

**Figure 7.**
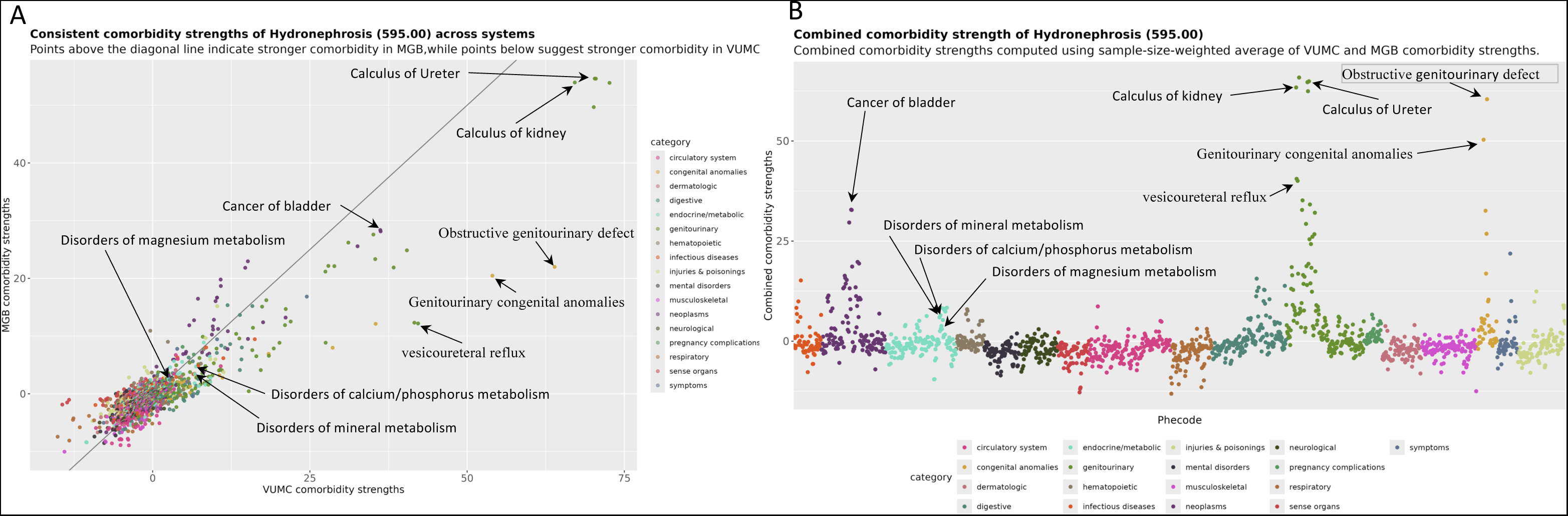
Multimorbidity Landscape of Hydronephrosis. **A.** Analysis reveals a largely consistent comorbidity landscape for hydronephrosis across both EHR systems. **A.** Pairwise comorbidity strengths are highly aligned, with minor variations in the extreme end of the distribution (e.g., obstructive genitourinary defect being slightly more comorbid in the Vanderbilt dataset). **B.** Consistent Phecode Comorbidity Strength identifies highly comorbid conditions, including expected associations such as obstructing stones, vesicoureteral reflux, congenital anomalies along with rarer causes and potentially novel conditions.

## Discussion

Our study demonstrates the robustness of EHR-derived multimorbidity analysis for investigating complex disease interrelations. We observed remarkable consistency in disease frequencies, comorbidity strengths, and multimorbidity network topologies across two large-scale, geographically distinct EHR systems. These findings underscore the potential of EHR-based phenome-wide analysis to uncover disease-disease interactions and clinically relevant clusters, despite variations in patient populations and healthcare practices.

The high concordance of multimorbidity patterns, after adjustment for potential confounders, provides compelling evidence for the reliability of this approach. Furthermore, the consistent core-periphery structure observed across networks offers a strategic framework for analysis, suggesting disease clusters within the core may share common etiologies or risk factors, while peripheral diseases may represent more specific mechanisms unique to subsets of patient population.

Notably, the alignment of multimorbidity patterns with genetic correlations further supports the potential of multimorbidity networks to unravel shared disease etiologies.

While multimorbidity patterns were largely conserved, we also observed nuanced differences that likely reflect demographic and regional variations among patient populations, as well as specialty differences across systems. This emphasizes the importance of contextualizing multimorbidity analyses within the specific framework of each EHR system, considering factors like demographics, age distribution, and regional health trends.

The use of phecodes as a disease phenome warrants specific consideration. Phecodes provide a standardized representation by grouping related ICD codes into broader, clinically relevant categories. Our results demonstrate that phecode-based multimorbidity analysis effectively captures reproducible disease relationships, supporting their use in large-scale EHR studies for consistent and transferable insights into comprehensive phenome-wide disease-disease relationships.

This study lays the methodological groundwork for advancing multimorbidity research. By identifying essential properties of multimorbidity networks derived from EHR data, such as consistent structural equivalence and a robust core-periphery structure, we provide promising directions for more sophisticated modeling approaches. For instance, probabilistic block modeling could be an effective approach for identifying disease clusters and quantifying patient enrichment with greater sensitivity and specificity. Our case study on hydronephrosis demonstrates the potential of multimorbidity networks in uncovering causal disease relationships, as illustrated by the replication of established causative disease associations. Future studies should develop advanced causal discovery models to quantify these relationships in depth, elucidating underlying mechanisms such as shared genetic or environmental predispositions. These insights are crucial for understanding the complex etiology of multimorbidity. Furthermore, robust multimorbidity networks offer a promising approach to identifying "controls" or "unexposed" patients with similar multimorbidity profiles to "cases" in observational studies, potentially improving the accuracy of traditional matching methods like propensity score matching.

Several limitations and future directions warrant consideration. While our pairwise comorbidity analysis provides valuable insights, it is only an initial step. Incorporating temporal disease progression patterns and diverse data sources (e.g., laboratory results, medications) could refine our understanding of disease trajectories and causal relationships. Additionally, while our null model simulations suggest minimal impact of phecode taxonomy structure on network conservation (Supplementary A15), a more in-depth analysis of how phenotype grouping methods affect findings warrants further investigation.

Overall, this study provides compelling evidence for the reproducibility and applicability of EHR-based multimorbidity network analysis, laying the groundwork for data-driven discoveries in precision medicine that aim to tailor prevention and treatment strategies to individual multimorbidity profiles. We have also introduced a knowledge base integrating multi-source data (Supplementary A18), serving as a resource for the research community to explore multimorbidity these complex relationships, fostering collaborations and accelerating discoveries in the field.

## Contributors

NS and YX conceived the idea and designed the study, DR offered significant guidance in both the design of the study and the data collection process. NS, TV, KC, YL and SZ primarily conducted the major statistical analysis and NS and SZ played a key role in the design and development of the web application. TV conducted data analysis and contributed significantly to the interpretation of the genetic data analysis. BS, QSH, EJP, JMP, CHW, MRS, YS, DMR were involved in data interpretation and preparation. NS and TV prepared the initial draft of the manuscript, and YX led revisions. RSH, CAB, LEW, JMB, DBJ, WHS, QC, TH, JWS, prepared and provided a critical revision of the manuscript for important intellectual content. All authors reviewed, provided valuable feedback, and gave their final approval for the published version of the manuscript.

## Data Sharing

All coding details associated with the models has been shared. Results have been aggregated and reported within this Article to the maximum extent possible, while maintaining privacy from personal health information as required by law. All dynamic online analysis results are available from PheMIME App (https://prod.tbilab.org/PheMIME/). All data are archived within TBILab systems in an audited computing environment secured by the Health Insurance Portability and Accountability Act to facilitate verification of study conclusions. The open-source code for PheMIME is publicly available on our GitHub repository at https://github.com/tbilab/PheMIME.

## Acknowledgments

NS and YX are supported by the Vanderbilt University Department of Biostatistics Development Award; YX, CB and RH are supported by R21DK127075; YX, DE, EP and DR are supported by P50GM115305; JWS is supported in part by R01 MH118233. The Vanderbilt University Medical Center dataset(s) used for the analyses described were obtained from Vanderbilt University Medical Center’s SD/BioVU, which is supported by numerous sources: institutional funding, private agencies, and federal grants. These include the NIH funded Shared Instrumentation Grant S10RR025141; and CTSA grants UL1TR002243, UL1TR000445, and UL1RR024975. Genomic data are also supported by investigator-led projects that include U01HG004798, R01NS032830, RC2GM092618, P50GM115305, U01HG006378, U19HL065962, R01HD074711; and additional funding sources listed at https://victr.vanderbilt.edu/pub/biovu/. This research has been conducted using the UK Biobank Resource under Application Number 43397.

