## Supplementary Materials for "Interoperability of phenome-wide multimorbidity patterns: a comparative study of two large-scale EHR systems"

#### A1. Dural regression analysis of disease comorbidities and pairwise disease comorbidity strength

Comorbidity strength reflects the association strength of one disease (phecode) occurring given the presence of another. It is used as the edge weight in the multimorbidity networks. In our analysis, two logistic regressions were run for every pair of phecodes, e.g., phecodes A, B. One modeled the probability of one phecode (A) occurring given the presence of another phecode (B), and vice versa. Both models were adjusted for patient demographics (age, sex, race) and disease burden (number of unique phecodes) to account for potential confounding factors that might influence the co-occurrence of diseases. The full models are specified as follows:

**For phecode B given A:**

$$\log\left(\frac{P(B|A, \text{covariates})}{1-P(B|A, \text{covariates})}\right) = \beta_{0B} + \beta_{1B}A + \gamma_{age,B}age + \gamma_{sex,B}sex + \gamma_{race,B}race + \gamma_{EHR\ age,B}EHR\ age + \gamma_{\log(\ disease\ burden)\ ,B}\ \log(\ disease\ burden) \qquad \text{(S1)}$$

**For phecode A given B:**

$$\log\left(\frac{P(A|B, \text{covariates})}{1-P(A|B, \text{covariates})}\right) = \beta_{0A} + \beta_{1A}B + \gamma_{age,A}age + \gamma_{sex,A}sex + \gamma_{race,A}race + \gamma_{EHR\ age,A}EHR\ age + \gamma_{\log(\ disease\ burden),A}\ \log(\ disease\ burden) \qquad \text{(S2)}$$

The Wald statistic was employed as a quantitative metric to evaluate the strength of association between phecodes. Rather than determining statistical significance, we used this measure to quantify the magnitude of association, or comorbidity strength, between pairs of phecodes. We computed the Wald statistics for the coefficient ($\beta_{1B}$ in mode 1 and $\beta_{1A}$ for the model 2) corresponding to the phecode of interest (B in model 1 and A in model 2) from each logistic regression model (S1 and S2, respectively), as follows:

$$Z_B = \frac{\beta_{1B}}{SE(\beta_{1B})}$$

$$Z_A = \frac{\beta_{1A}}{SE(\beta_{1A})}$$

To address the directional nature of logistic regression, we averaged the Wald statistics, $Z_A$ and $Z_B$, and formulate a symmetrical score, which we designate as the Pairwise Comorbidity Strength (PCS) between the two phecodes. The PCS for a pair of phecodes, A and B, is computed as follows:

$$PCS_{AB} = \frac{Z_A + Z_B}{2}$$

We conducted our analysis in R version 4.3.2. Specifically, we applied the `glm()` function with `family = binomial` for the logistic regressions. From the model summaries, we extracted the $\beta$ coefficients and their standard errors, computed the Wald statistics and the Pairwise Comorbidity Strength (PCS) scores for each phecode pairs.

#### A2. Quantify phecode similarity and conservation in multimorbidity networks using structural equivalence

Structural equivalence is a concept used to identify nodes that exhibit similar patterns of connections to other nodes in the network. Nodes are considered structurally equivalent if they share common neighbors within a network(Lorrain & White, 1977; Newman, 2018). As depicted schematically in Figure S1, nodes A and B demonstrate structural equivalence by sharing common neighbors X, Y, and Z. Perfect structural equivalence is uncommon in real networks, various metrics have been developed to quantify the degree of equivalence or similarity between the connection patterns of the node. A widely used metric is Pearson's correlation coefficients, which measures the similarity between the connection profiles of two nodes, taking into account the quantitative weights of the edges.

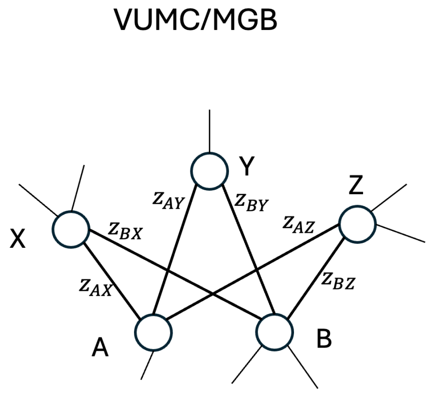

**Figure S1. Structural equivalence and phecode similarity.**

Our multimorbidity network is an undirected, weighted network, where nodes are diseases (phecodes) and edges represent disease-disease connections weighted by their comorbidity strengths, derived from the dual regression analysis. For each disease pair, say A and B, we calculated Pearson's correlation coefficient between their respective weighted adjacency vectors (i.e., multimorbidity patterns), denote as $\boldsymbol{Z}_A = (Z_{1A}, Z_{2A}, \dots, Z_{nA})'$ and $\boldsymbol{Z}_B = (Z_{1B}, Z_{2B}, \dots, Z_{nB})'$. The resulting coefficient, termed $Similarity_{AB}$, quantifies the linear correlation between the multimorbidity patterns of A and B, defined based on their comorbidity strengths with other diseases. Specifically, $Similarity_{AB}$ can be calculated using:

$$Similarity_{AB} = \frac{\sum_i (Z_{iA} - \bar{Z}_A)(Z_{iB} - \bar{Z}_B)}{\sqrt{\sum_i (Z_{iA} - \bar{Z}_A)^2}\sqrt{\sum_i (Z_{iB} - \bar{Z}_B)^2}}, \qquad i = 1, \dots 1815 \text{ (Phecode } v1.2)$$

Here, $Z_{iA}$ and $Z_{iB}$ represent comorbidity strengths of diseases A and B with a third disease *i*, respectively, and $\bar{Z}_A$ denotes the average pairwise comorbidity strength of Phecode A with all other phecodes, corresponding to the mean value of the element values of $\boldsymbol{Z}_A$. Similarly, $\bar{Z}_B$ denotes the average comorbidity strength for phecode B with all other phecodes, calculated as the mean of the element values in $\boldsymbol{Z}_B$.

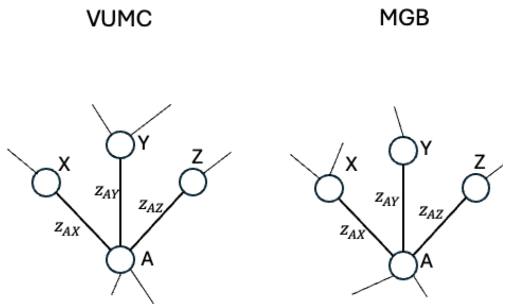

**Figure S2. Structure equivalence and phecode conservation.**

In our study, we used consistent phecode mappings (Phecode v1.2) across both the Vanderbilt University Medical Center (VUMC) and the Massachusetts General Brigham (MGB) EHR systems. This allows us to directly compare the structural equivalence of phecodes and assessing the conservation (or variability) of their multimorbidity patterns across systems. Figure S2 provides a schematic illustration of the concept of phecode conservation. To quantify conservation in our multimorbidity networks, we calculated Pearson's correlation coefficient between a phecode's (A's) multimorbidity patterns in VUMC (V) and MGB (M), denote as $\boldsymbol{Z}_A^V = (Z_{1A}^V, Z_{2A}^V, \ldots, Z_{nA}^V)'$ and $\boldsymbol{Z}_A^M = (Z_{1A}^M, Z_{2A}^M, \ldots, Z_{nA}^M)'$, respectively. Coefficient values close to 1 indicate strong conservation, suggesting the phecode exhibits a similar pattern of comorbidities with other diseases across both systems. Specifically, we calculated the conservation of Phecode A between VUMC and MGB, denoted as $Conservation^{V,M}{}_A$, using the following formula:

$$Conservation_A^{(V,M)} = \frac{\sum_i (Z_{iA}^V - \bar{Z}^V{}_A)(Z_{iA}^M - \bar{Z}^M{}_A)}{\sqrt{\sum_i (Z_{iA}^V - \bar{Z}^V{}_A)^2}\sqrt{\sum_i (Z_{iA}^M - \bar{Z}^M{}_A)^2}}, \qquad i = 1, \ldots 1815 \text{ (Phecode } v1.2)$$

where, $Z_{iA}^V$ and $Z_{iA}^M$ represent comorbidity strengths between phecodes A with phecode *i* in the VUMC (V) and MGB (M) systems, respectively. Additionally, and $\bar{Z}^V{}_A$ denotes the average pairwise comorbidity strength of Phecode A with all other phecodes in the VUMC system (i.e. mean of the element values of $\boldsymbol{Z}_A^V$), while $\bar{Z}^M{}_A$ represents the corresponding average of $\boldsymbol{Z}_A^V$ for the MGB system.

#### A3. Spectral analysis and global and meso-scale structures of multimorbidity networks

Spectral analysis is a useful tool for investigating complex network structures at various scales. It involves studying the eigenvalues and eigenvectors of a network's adjacency matrix. Eigenvalues reveal information about overall network connectivity, while the leading eigenvector (corresponding to the largest eigenvalue) often provides insights into node centrality and importance within the network(Cvetkovic et al., 1997).

We assessed the global-scale topological consistency of multimorbidity networks across institutions using leading eigenvector centrality, which quantifies a disease node's importance based on the number, strength, and importance of its connections with other diseases. Analyzing and comparing the distribution of the centrality across two systems reveals overall network structure and its consistency. Our analysis revealed a high correlation in eigenvector centralities between VUMC and MGB (Spearman correlation = 0.902; 95% CI: 0.889 – 0.914, Figure 3A), indicating a consistent global network structure of multimorbidity networks.

To evaluate the meso-scale structure of the multimorbidity networks, we applied the eigengap heuristic(Schoch et al., 2017). This metric is calculated as $1 - \lambda_2/\lambda_1$, where $\lambda_1$ and $\lambda_2$ are the largest and second-largest eigenvalues of the network adjacency matrix (A), which was constructed using a p-value cutoff of 0.05. (Note that if the more stringent Bonferroni cutoff of 0.05 divided by the number of tests were applied, the eigengap values would be 0.71 and 0.68, respectively). In networks with a clear core-periphery structure, the largest eigenvalue ($\lambda_1$ ) is significantly larger than the second ($\lambda_2$), which leads to a high eigengap value. A higher eigengap value indicates a stronger core-periphery structure, with values close to 1 suggesting a near-ideal core-periphery structure – a threshold graph. Our multimorbidity networks exhibited a strong core-periphery structure, as evidenced by consistently high eigengap values across VUMC and MGB (0.836 and 0.844 respectively). In addition, the uniformly high eigengap values across systems indicate both the presence of a strong core-periphery structure and its consistency across the two systems.

For threshold graphs, common centrality indices such as degree, closeness, betweenness, and subgraph centrality are all substantially correlated due to the neighborhood-inclusion preorder property (SCHOCH & BRANDES, 2016). As depicted in Figure S3, if node B in the network is connected to all the neighbors of node A (and possibly more), then B has a more inclusive neighborhood than A. This establishes a hierarchy where nodes with more inclusive neighborhoods consistently have higher centrality scores across all these measures. When centrality measures are consistent across systems, this hierarchy becomes an inter-system feature, suggesting that the underlying network structure is more important than the nuances between different centrality metrics. In this case, eigenvector centrality serves as a representative measure of this structural invariant of the network (Oldham et al., 2019; Opsahl et al., 2010; Schoch et al., 2017).

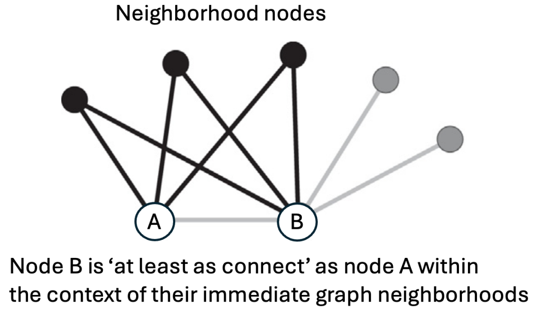

**Figure S3. Neighborhood inclusion preorder: B is dominated by A**

Our multimorbidity networks closely resemble threshold graphs, as indicated by the consistent high eigengap values across systems. This suggests the potential presence of a neighborhood inclusion preorder in multimorbidity networks, with interesting implications for both the networks themselves and their consistency across institutions: (1) If the neighborhood inclusion preorder holds true, it implies a predictable ranking of diseases based on their centrality measures (degree, closeness, betweenness, etc.). Diseases with more inclusive neighborhoods (connected to a broader range of other diseases) will consistently have higher centrality scores across all these measures. This ranking could help identify "central" diseases that play a crucial role in the overall disease network structure. (2) High correlations of centrality measures across the systems might indicate the presence of tightly connected disease clusters with consistent relative positions across different EHR systems. These clusters could represent groups of diseases with strong co-occurrence patterns or shared essential underlying biological mechanisms. Investigating such clusters could provide valuable insights into disease relationships. (3) Understanding the centrality ranking and potential disease clusters can inform disease prioritization within the network. Diseases with high centrality scores (and potentially central roles in clusters) might be more critical targets for interventions that aim to improve the overall efficacy of the healthcare system, optimize patient management, and outcomes. In summary, the potential presence of a neighborhood inclusion preorder adds another layer of consistency analysis to your multimorbidity networks. While further investigation is needed to confirm its properties, it offers valuable insights into disease importance, clustering patterns, and network robustness.

We utilized the R package igraph v2.0.0 to analyze the multimorbidity networks. First multimorbidity networks were created using graph_from_data_frame(). Then, we convert the network into its adjacency matrix using the function `as_adjacency_matrix()`. Finally, we calculate the eigenvalues of the adjacency matrix using the base R function `eigen()`, which returns the eigenvalues as a vector. We then calculated eigenvector centrality scores for each node in the network using the `eigen_centrality()` function, which returns a vector containing the centrality scores. We also calculated eigengap values using the eigenvalues.

#### A4. Threshold graph construction and implication to cluster analysis of multimorbidity networks

Threshold graph is a special type of graph with a simple, yet structured, construction process. Two formal definitions exist: (1) A threshold graph is a graph G=(V, E), where V is the set of vertices (nodes) and E is the set of edges, that can be constructed from an empty graph by repeatedly adding either an isolated vertex (not connected to any others) or a dominating vertex (connected to all existing vertices). This sequential construction ensures uniqueness up to isomorphism, meaning any two threshold graphs built from the same rules are essentially the same, just with different vertex labels. (2) Alternatively, threshold graphs can be defined using a real-valued threshold (T) and a weight function (w) that assigns weights to vertices (w: V→ R, where R is the set of real numbers). For any two distinct nodes u, v ∈ V, an edge e = {u, v} ∈ E exists if and only if the sum of their weights exceeds T: w(u) + w(v) > T

These definitions have profound implications for multimorbidity network analysis: (1) The threshold-based definition aligns well with multimorbidity networks. We can use comorbidity strength values as node weights. For diseases A and B, their weights are defined as:

$$w_B = \frac{Z_B}{2} = \frac{\beta_{1B}}{2SE(\beta_{1B})}$$

$$w_A = \frac{Z_A}{2} = \frac{\beta_{1A}}{2SE(\beta_{1A})}$$

An edge (connection) exists between A and B if their combined weight exceeds a chosen threshold ($w_B$+$w_A$ >T). This is equivalent to their pairwise comorbidity score ($PCS_{AB} = w_B + w_A$) exceeding the threshold. (2) The weighted representation of threshold graphs naturally induces a hierarchical clustering on the network. Diseases with strong comorbidity (high pairwise comorbidity strength value) could be isolated and clustered at higher thresholds, while weaker comorbidities emerge at lower thresholds. This allows us to systematically investigate disease clusters at different cutoff levels of comorbidity strength. (3) Threshold graphs are unique up to isomorphism. Sequential construction of multimorbidity networks provide a consistent solution to represent the network unique up to isomorphism. Together with the consistent structural equivalence across the systems, the hierarchical cluster analyses are likely to identify consistent and reproductible diseases clusters across the systems. Overall, the properties of threshold graphs offer powerful theoretical lens through which to analyze and understand the complex patterns of multimorbidity networks, leading to robust and insightful analyses, and potentially revealing consistent biological mechanisms or shared risk factors underlying multiple diseases.

#### A5. Dynamic network clustering of multimorbidity networks using *AssociationSubgraphs*

We employed the AssociationSubgraphs algorithm (Strayer et al., 2023), an interactive clustering approach, to identify the subgraphs (disease clusters) within multimorbidity networks. Inspired by threshold graph construction, this algorithm focuses on discovering clusters of diseases with strong comorbid relationships. It begins by sorting network edges (representing disease-disease connections) in descending order of comorbidity strength. The nodes connected by the edge with the highest comorbidity strength form the initial cluster. Subsequently, edges are added iteratively; if one node belongs to an existing cluster, the other node joins that cluster. If both nodes are unconnected, they form a new cluster, and if they belong to separate existing clusters, those clusters are merged. The algorithm exports the cluster state after each edge addition, allowing us to track cluster evolution. To determine the optimal cutoff point, we utilize the 'largest-smallest' rule (Strayer et al., 2023), which pinpoints the transition from well-defined clusters to the emergence of a giant component where randomness begins to dominate. For a comprehensive explanation of the algorithm, please refer to Strayer et al., 2023..

The AssociationSubgraphs algorithm leverages the properties of the threshold graph and implements a dynamic network clustering strategy, identifying subgraphs at all unique values of comorbidity strength. By tracking how subgraphs evolve as edges are added in descending order of comorbidity strength, the algorithm provides insights into the hierarchical structure of disease clusters and their merging patterns at different levels of comorbidity strength. The threshold process and the hierarchical structure on the identified disease clusters allows for the identification of broader, clinically relevant clusters, even when diseases within a cluster do not exhibit dense connectivity or uniformly strong pairwise comorbidities – a scenario where traditional community detection methods might struggle.

#### A6. Comparison of phecode frequencies between VUMC and MGB

Consistent disease frequencies provide the informational foundation for the robustness of ICD-based disease frequencies, which provided the foundational information for the assessment of covariate-adjusted comorbidity strength, as well as the consistency of multimorbidity networks.

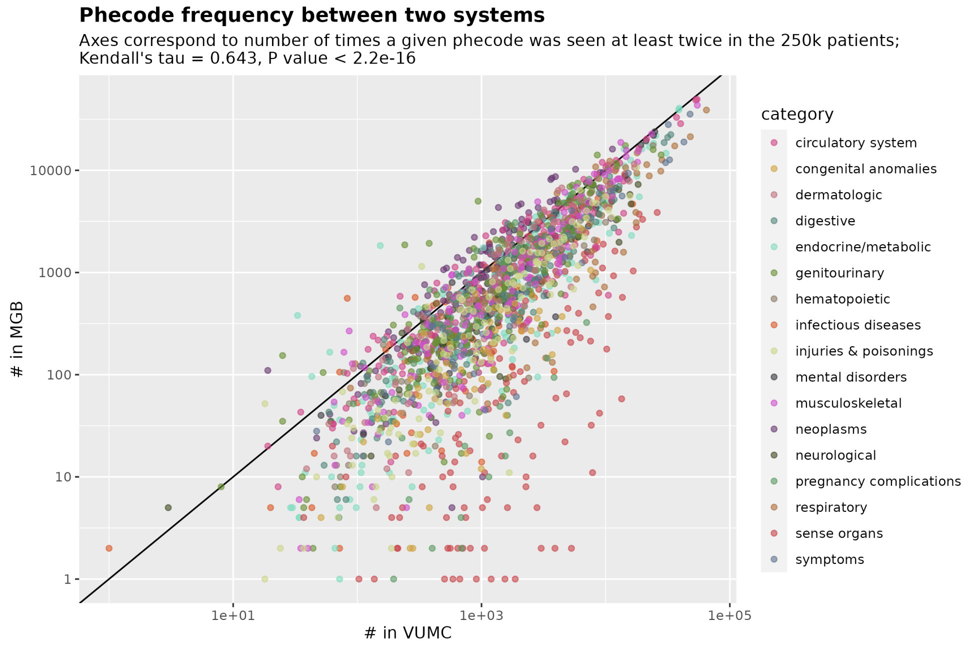

**Figure S4. Correlation of Phecode Frequencies.** Phecode frequencies across VUMC and MGB show high consistency (Kendall's $\tau = 0.643$).

#### A7. Phecode multimorbidity conservation by category

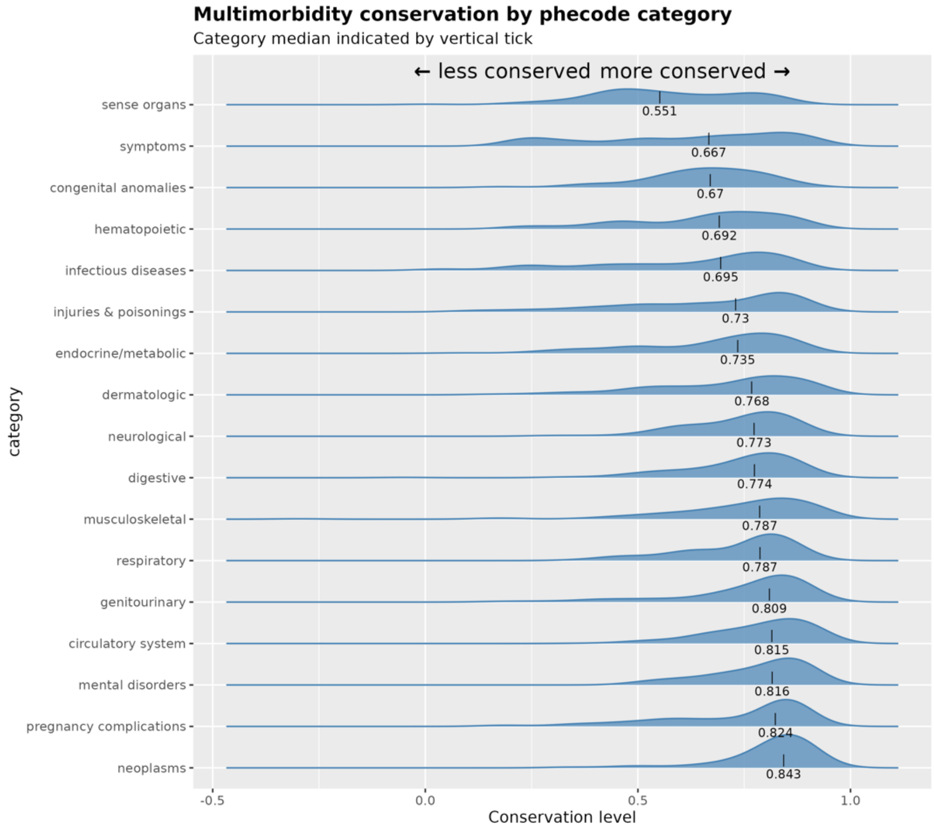

**Figure S5. Conservation of Phecode Multimorbidity Patterns.** The distribution of phecode multimorbidity conservation by disease category are skewed toward 1, showing a strong conservation of phecode multimorbidity patterns.

### A8. Most and least conserved phecodes multimorbidity patterns

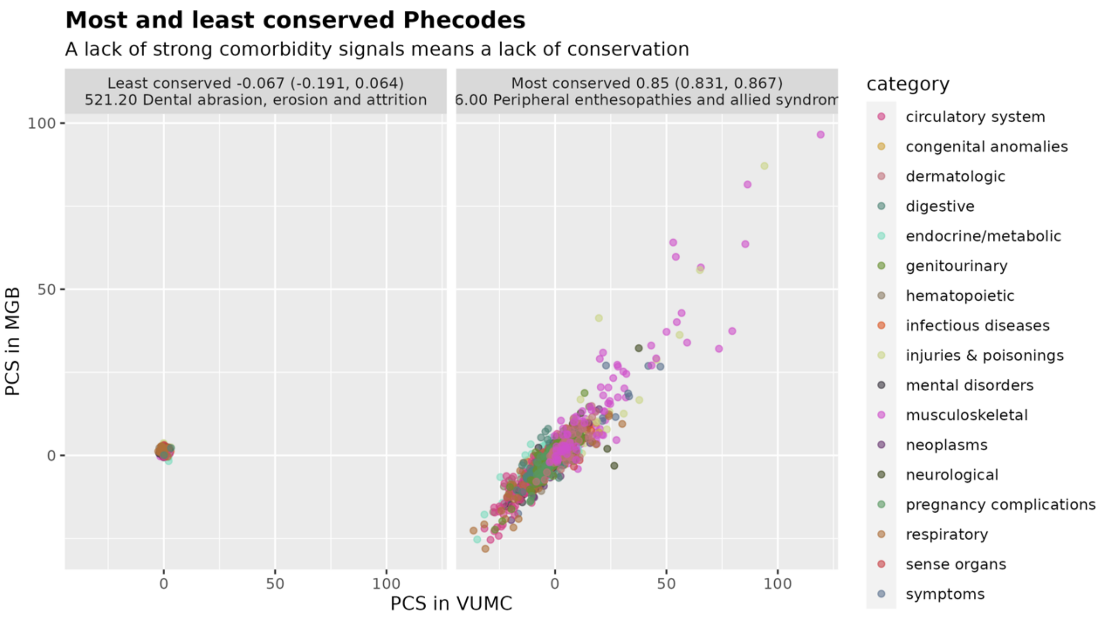

**Figure S6. Pairwise Comorbidity Strength (PCS) in Most Conserved vs. Least Conserved Phecodes.** Scatterplot reveals a stronger positive correlation between PCS and conservation level in the most conserved phecodes (right), compared to the least conserved phecodes (left).

#### A9. Distributions of centrality differences by category

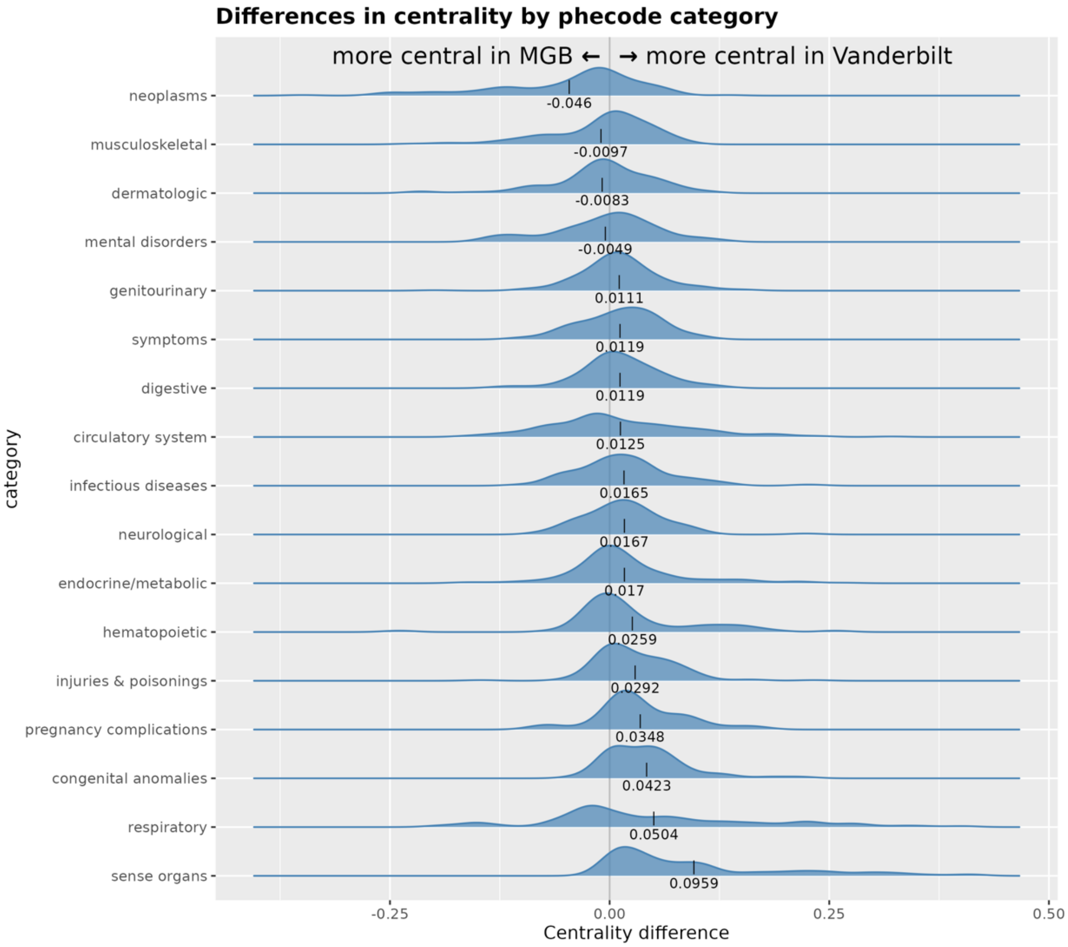

**Figure S7. Distribution of Phecode Centrality Differences Across Systems.** The centrality differences for each phecode category predominantly cluster around zero, suggesting a high degree of consistency between systems.

#### A10. A combined multimorbidity network

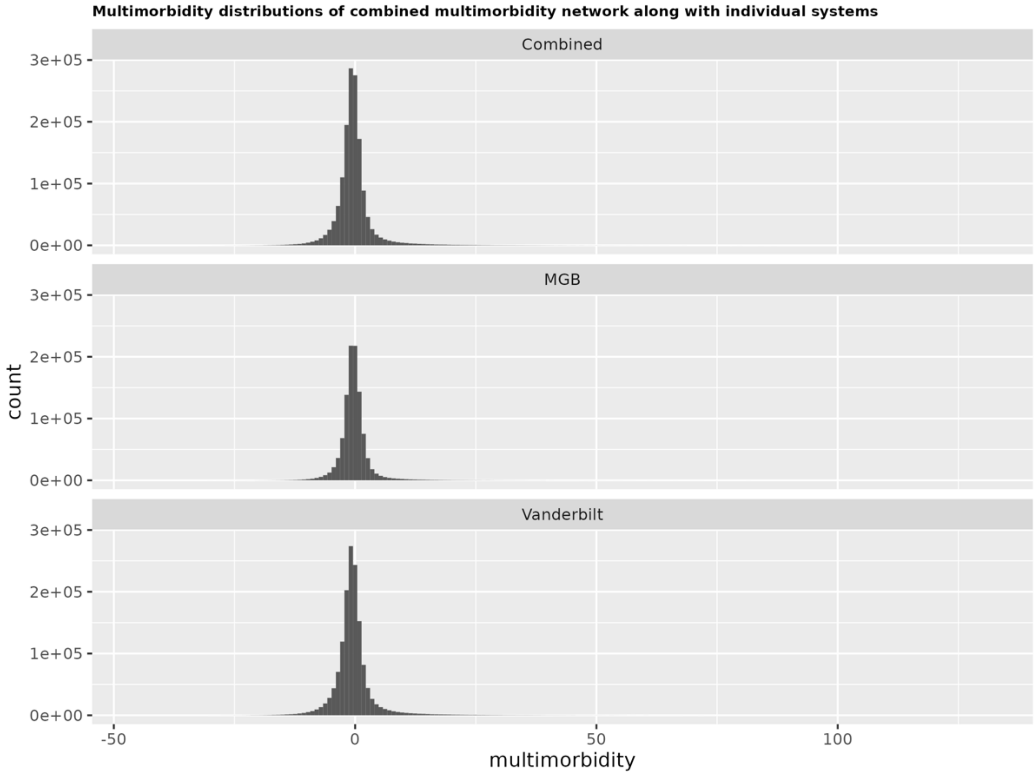

**Figure S8. Combined Multimorbidity Network and Distribution.** This figure illustrates the network formed using a weighted average of comorbidity strengths from two systems. Phecode pair associations are weighted by the number of patients exhibiting the comorbidity in each system. The resulting combined comorbidity strength distribution maintains a Gaussian shape, as expected from the product of two Gaussian distributions.

#### A11. UMPA projection of multimorbidity similarity networks

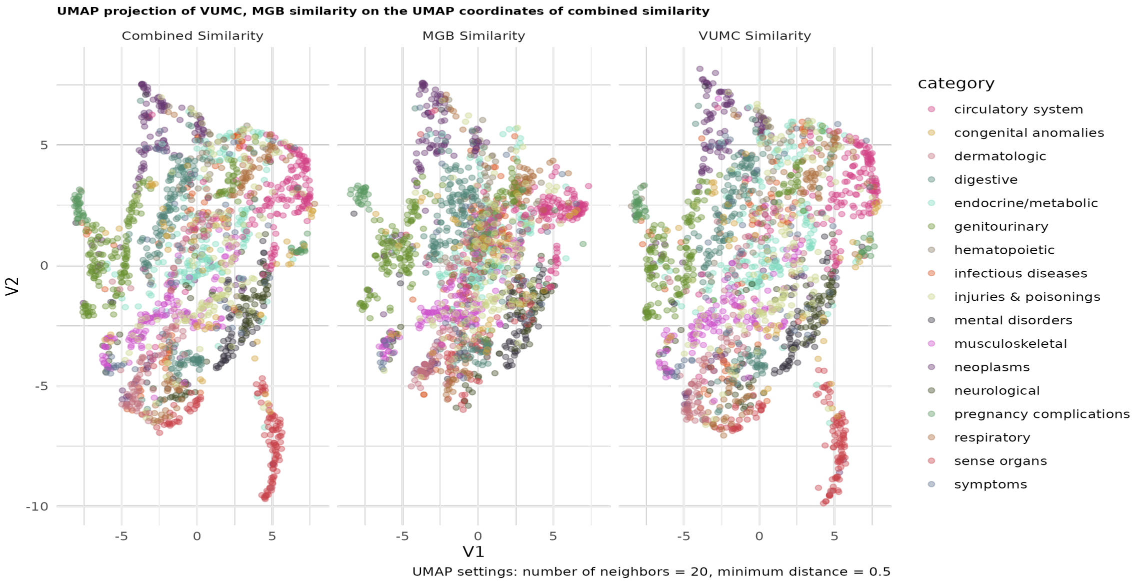

**Figure S9. Multimorbidity Similarity Networks with UMAP Visualization.** This figure demonstrates the clustering of phecodes based on their multimorbidity patterns. UMAP dimension reduction reveals distinct groupings of phecodes within their respective categories, as well as potential subclusters deserving further investigation.

#### A12. Top phecodes with highest centrality measurements

**Table S3. Phecodes with Highest Centrality in VUMC and MGB Comorbidity Networks.** This table highlights the phecodes exhibiting the highest centrality measures within each network. Sense organ and respiratory phecodes demonstrate greater centrality in the VUMC network, while neoplasms, musculoskeletal, and dermatologic phecodes are more central in the MGB network. Centrality is measured using normalized eigenvector centrality within the symmetric comorbidity association network, with the most central node having a centrality score of 1.

| | | Centrality* | | |
|---|---|---|---|---|
| | Category | Vandy | MGB | Difference† |
| **More central in Vanderbilt** | | | | |
| 366.00 Cataract | sense organs | 0.538 | 0.127 | 0.412 |
| 381.00 Otitis media and Eustachian tube disorders | sense organs | 0.726 | 0.317 | 0.409 |
| 464.00 Acute sinusitis | respiratory | 0.748 | 0.346 | 0.402 |
| 381.20 Eustachian tube disorders | sense organs | 0.515 | 0.146 | 0.369 |
| 476.00 Allergic rhinitis | respiratory | 0.867 | 0.513 | 0.355 |
| 371.00 Inflammation of the eye | sense organs | 0.489 | 0.144 | 0.344 |
| 483.00 Acute bronchitis and bronchiolitis | respiratory | 0.593 | 0.265 | 0.329 |
| 379.00 Other disorders of eye | sense organs | 0.398 | 0.071 | 0.327 |
| 366.20 Senile cataract | sense organs | 0.383 | 0.059 | 0.324 |
| 414.00 Other forms of chronic heart disease | circulatory system | 0.661 | 0.340 | 0.321 |
| **More central in MGH** | | | | |
| 694.20 Other dyschromia | dermatologic | 0.417 | 0.633 | −0.216 |
| 198.40 Secondary malignant neoplasm of liver | neoplasms | 0.324 | 0.552 | −0.228 |
| 740.90 Osteoarthrosis NOS | musculoskeletal | 0.529 | 0.758 | −0.228 |
| 289.40 Lymphadenitis | hematopoietic | 0.348 | 0.587 | −0.239 |
| 198.20 Secondary malignancy of respiratory organs | neoplasms | 0.380 | 0.621 | −0.242 |
| 198.30 Secondary malignant neoplasm of digestive systems | neoplasms | 0.302 | 0.554 | −0.252 |
| 165.10 Cancer of bronchus; lung | neoplasms | 0.320 | 0.574 | −0.254 |
| 196.00 Radiotherapy | neoplasms | 0.281 | 0.536 | −0.255 |
| 165.00 Cancer within the respiratory system | neoplasms | 0.327 | 0.585 | −0.258 |
| 198.00 Secondary malignant neoplasm | neoplasms | 0.589 | 0.939 | −0.350 |

* Centrality is normalized eigen-centrality of node given symmetric comorbidity associations. Most central node has eigen-centrality of 1.

† Difference = $\text{Centrality}_{\text{Vandy}}$ - $\text{Centrality}_{\text{MGB}}$

#### A13. Genotype sample description, ancestry estimation, and quality control (QC)

The VUMC Samples were genotyped on the Infinium expanded multi-ethnic genotyping array (MEGAEx)27 from Vanderbilt's biobank, BioVU, which initially contained 2,038,233 single nucleotide polymorphisms (SNPs) and 94,474 people. At first, we excluded the SNPs with missing rate >= 0.05 and the positional duplicate SNPs with incompatible alleles. Then we excluded individuals with call rate < 98%, sex discordance, excess heterozygosity rate within each self-reported ancestry, or potentially cross-contaminated individuals [proportion identity-by-descent (IBD) > 0.8 between different individual IDs]. We next excluded SNPs with missing rate > 0.02, minor allele frequency (MAF) < 0.01, Hardy-Weinberg equilibrium (HWE) test P value ≤ 1x10–6 within self-reported ancestry of AFR or EUR, or MAF < 0.01 within the whole sample. Thus, there were 90,313 individuals which were kept with a set of high-quality autosomal SNPs (N=887,250).

We downloaded 1000 Genomes phase 3 (1000 GP3) data28, which consists of 2,504 unrelated samples from 5 super populations African (AFR), Admixed American (AMR), East Asian (EAS), European (EUR), South Asian (SAS). Next, we extracted 887,250 QC-ed SNPs (no INDELs) that are genotyped on the BioVU MEGAEx array and merged it with 1000 GP3 data after removing C/G and A/T SNPs to avoid unresolvable strand mismatches in MEGA samples. Regions with known high linkage disequilibrium (LD) were excluded and the common variants were then pruned (r2 < 0.05) using PLINK 1.9 (–indep-pairwise 1000 50 0.05) to yield 71,339 SNPs in relative linkage equilibrium for ancestry analyses.29 The flashpca version 2.0 was used to perform PCA to generate 2 top genetic PCs.30 By using K nearest neighbors (KNN) clustering with k = 5, we inferred MEGA samples' ancestries based on the top 2 PCs from the 1000 GP3 and MEGA samples. We treated 1000 GP3 samples as the training set and MEGA samples as the testing set. For each MEGA individual, we calculated its Euclidean distance from each individual in the training set. Next, we sorted the distance, and found the 5 nearest neighbors from the 1000 GP3 based on the 5-th minimum distances for each MEGA individual. Based on whether all of the nearest neighbors were from the same population, we inferred the detected MEGA individual's ancestry. (If they are from more than one super population, we assigned the individual's ancestry as admixed one.) Among the 90,313 MEGA individuals, 87,558 (96.95%) were assigned to five homogeneous super-populations, i.e. AFR=13,808, AMR=2,446, EUR=70,473, EAS=441, SAS=390.31 The final set used in our main analyses was the EUR only subgroup.

Prior to imputation, the SNP position, alleles, Ref/Alt assignments and frequency differences were checked by comparison with the Haplotype Reference Consortium (HRC) panel (Version r1.1 2016)32 in build GRCh3733 using McCarthy Group Tools (https://www.well.ox.ac.uk/~wrayner/tools/). SNPs with inconsistent alleles, with > 0.2 allele frequency difference, or not in the reference panel were removed. Phasing and imputation were then performed using a standard pipeline on the Michigan Imputation Server (MIS).34 The total QC-ed SNP data was divided into five batches. Phasing was performed using Eagle version v2.4.32 The HRC reference panel in build GRCh37 was selected using mixed population.32,33 The genotype probabilities in post-imputed data were converted to hard-call genotypes using PLINK2 (hard-call >= 0.1).35 SNPs were removed with imputation info score in any of the batches < 0.3, position duplicates, missing genotype rate > 0.02, or multi-allelic states (>2). For autosomal data, we reused a series of QC filtering steps, excluding SNPs with MAF < 0.005, HWE test P value < 1x10–6, and removing the individuals with missing rate ≥ 0.02, excess heterozygosity rate over 3* interquartile range (IQR) of the upper heterozygosity quartile (Q3) for each subset.

#### A14. Genetic correlation of 15 prevalent and heritable phenotypes

Understanding the genetic relationships between phenotypically similar diseases can provide valuable insights into their shared biological pathways and potential treatment targets. In this section, we investigate the genetic correlations of 15 prevalent, heritable phenotypes and explore their connections to observed multimorbidity patterns. The phenotypes were selected on being well studied, having at least 5,000 samples with at least 2 diagnostic codes and significant SNP-heritability within VUMC's biobank and GWAS summary statistics available from PheWeb.36 The list was further filtered to exclude uninteresting phecodes, ultimately leaving 15 phenotypes. GWAS summary statistics for each of the 15 phecodes (244.4, 250.2, 272.1, 278.1, 296.2, 300.1, 318, 401.1, 411.2, 411.4, 427.7, 495, 562.1, 585.1, 740.11) were obtained from the UK Biobank online tool PheWeb.36 The munge_sumstats.py script from the linkage disequilibrium score regression (LDSC) software version 1.0.1 was used to convert the summary statistics into the LDSC acceptable format while also removing poor quality SNPs.37 N for each phecode was determined by the number of cases and controls reported on PheWeb.

We used LDSC to calculate pairwise genetic correlations among the 15 phenotypes, using European Ancestry LD scores as a reference. The genetic correlation results are presented in a correlation plot (Figure S10). Additionally, we calculated the Pearson correlations between VUMC-based comorbidity strength and genetic correlation, as well as the correlations between VUMC-based multimorbidity similarity and genetic correlation for each phenotype pair (Figures 5A and 5B, respectively, and also in Figure S11 with corresponding disease pairs indexed so they can be looked up in the Table S4). The combined network metric was not used in these calculations due to its unique construction.

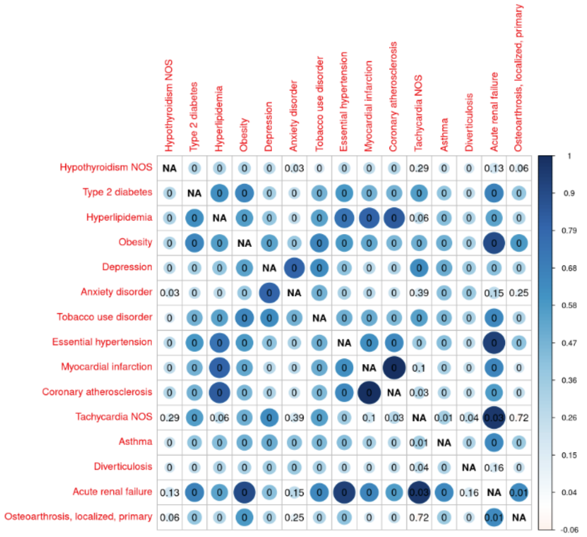

**Figure S10. Genetic Correlation of 15 Prevalent and Heritable Phenotypes**

a

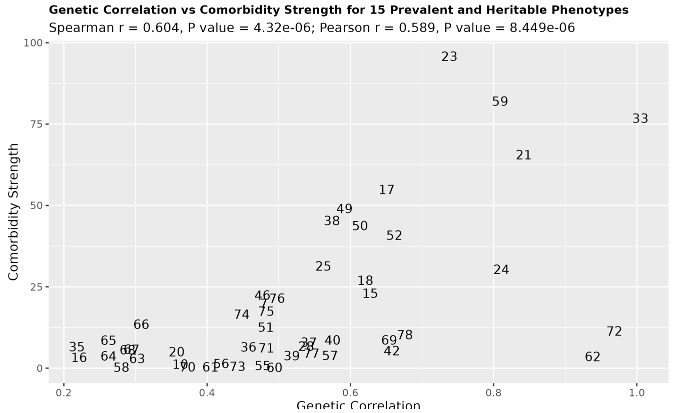

b

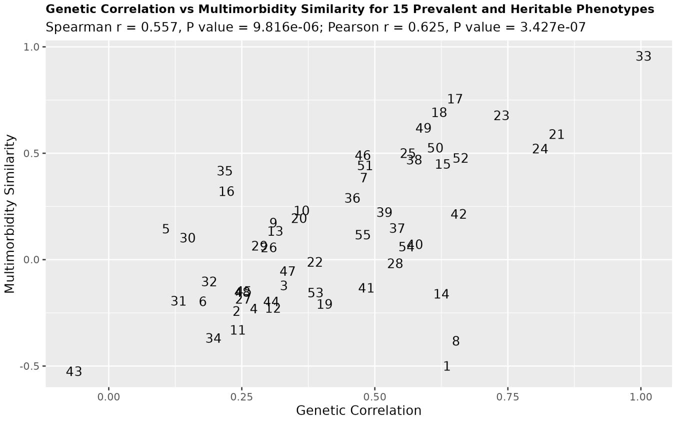

**Figure S11. Relationship between phenotypic similarity and genetic correlation.** Scatterplots demonstrate a positive association between both measures of phenotypic similarity (comorbidity strength and multimorbidity similarity) and genetic correlations across 15 heritable phenotypes. (a) VUMC comorbidity strength vs. genetic correlation. (b) Multimorbidity similarity vs. genetic correlation. **See Table S0 for phenotype pairs.**

**Table S4. Comorbidity strength, multimorbidity similarity, genetic correlations of 105 disease pairs across 15 heritable phenotypes.**

| Index | Phenotype a | Phenotype b | Genetic Correlation | Comorbidity Strength | Multimorbidity Similarity |
|---|---|---|---|---|---|
| 1 | Acute renal failure | Osteoarthrosis_localized_primary | 0.6358 | -13.559 | -0.498 |
| 2 | Anxiety disorder | Tobacco use disorder | 0.4793 | 19.9 | 0.392 |
| 3 | Anxiety disorder | Essential hypertension | 0.3291 | -0.492 | -0.167 |
| 4 | Anxiety disorder | Myocardial infarction | 0.2729 | -6.331 | -0.252 |
| 5 | Anxiety disorder | Coronary atherosclerosis | 0.2405 | -10.835 | -0.262 |
| 6 | Anxiety disorder | Tachycardia NOS | 0.1769 | -2.594 | -0.183 |
| 7 | Anxiety disorder | Asthma | 0.42 | 1.446 | 0.169 |
| 8 | Anxiety disorder | Diverticulosis | 0.3714 | -2.971 | 0.165 |
| 9 | Anxiety disorder | Acute renal failure | 0.2701 | -11.226 | -0.34 |
| 10 | Anxiety disorder | Osteoarthrosis_localized_primary | 0.1078 | -4.979 | 0.114 |
| 11 | Asthma | Diverticulosis | 0.3094 | -1.005 | 0.186 |
| 12 | Asthma | Acute renal failure | 0.6525 | -12.985 | -0.373 |
| 13 | Asthma | Osteoarthrosis_localized_primary | 0.3627 | 1.216 | 0.185 |
| 14 | Coronary atherosclerosis | Tachycardia NOS | 0.2805 | 0.243 | 0.49 |
| 15 | Coronary atherosclerosis | Asthma | 0.2988 | -5.468 | -0.134 |
| 16 | Coronary atherosclerosis | Diverticulosis | 0.2371 | -12.307 | -0.418 |
| 17 | Coronary atherosclerosis | Acute renal failure | 0.5718 | 3.941 | 0.476 |
| 18 | Coronary atherosclerosis | Osteoarthrosis_localized_primary | 0.2426 | -15.883 | -0.372 |
| 19 | Depression | Anxiety disorder | 0.8092 | 82.052 | 0.764 |
| 20 | Depression | Tobacco use disorder | 0.6278 | 23.061 | 0.454 |
| 21 | Depression | Essential hypertension | 0.4048 | 0.373 | -0.092 |
| 22 | Depression | Myocardial infarction | 0.2804 | -8.248 | -0.219 |
| 23 | Depression | Coronary atherosclerosis | 0.3089 | -14.323 | -0.24 |
| 24 | Depression | Tachycardia NOS | 0.6254 | -6.325 | -0.158 |
| 25 | Depression | Asthma | 0.4944 | 0.18 | 0.099 |
| 26 | Depression | Diverticulosis | 0.3474 | -3.66 | 0.109 |
| 27 | Depression | Acute renal failure | 0.4204 | -8.814 | -0.262 |
| 28 | Depression | Osteoarthrosis_localized_primary | 0.313 | -5.503 | 0.13 |

| | | | | | |
|---|---|---|---|---|---|
| 29 | Diverticulosis | Acute renal failure | 0.1776 | -6.943 | -0.36 |
| 30 | Diverticulosis | Osteoarthrosis_localized_primary | 0.2214 | 3.26 | 0.325 |
| 31 | Essential hypertension | Myocardial infarction | 0.6209 | 27.031 | 0.751 |
| 32 | Essential hypertension | Coronary atherosclerosis | 0.6509 | 54.89 | 0.801 |
| 33 | Essential hypertension | Tachycardia NOS | 0.3577 | 4.984 | 0.357 |
| 34 | Essential hypertension | Asthma | 0.4255 | -5.03 | -0.155 |
| 35 | Essential hypertension | Diverticulosis | 0.2683 | -4.141 | -0.271 |
| 36 | Essential hypertension | Acute renal failure | 0.9385 | 3.572 | 0.472 |
| 37 | Essential hypertension | Osteoarthrosis_localized_primary | 0.4057 | -10.241 | -0.277 |
| 38 | Hyperlipidemia | Obesity | 0.5625 | 31.41 | 0.508 |
| 39 | Hyperlipidemia | Depression | 0.3875 | -1.268 | -0.018 |
| 40 | Hyperlipidemia | Anxiety disorder | 0.2603 | -1.247 | -0.031 |
| 41 | Hyperlipidemia | Tobacco use disorder | 0.5384 | 6.713 | 0 |
| 42 | Hyperlipidemia | Essential hypertension | 0.7384 | 95.792 | 0.7 |
| 43 | Hyperlipidemia | Myocardial infarction | 0.811 | 30.351 | 0.535 |
| 44 | Hyperlipidemia | Coronary atherosclerosis | 0.8423 | 65.571 | 0.603 |
| 45 | Hyperlipidemia | Tachycardia NOS | 0.2528 | -13.992 | -0.061 |
| 46 | Hyperlipidemia | Asthma | 0.4236 | -4.854 | 0.063 |
| 47 | Hyperlipidemia | Diverticulosis | 0.3024 | 2.984 | -0.029 |
| 48 | Hyperlipidemia | Acute renal failure | 0.5329 | -11.493 | 0.085 |
| 49 | Hyperlipidemia | Osteoarthrosis_localized_primary | 0.3011 | -5.849 | -0.026 |
| 50 | Hypothyroidism NOS | Type 2 diabetes | 0.2891 | 5.611 | 0.286 |
| 51 | Hypothyroidism NOS | Hyperlipidemia | 0.3082 | 13.436 | 0.302 |
| 52 | Hypothyroidism NOS | Obesity | 0.2946 | 5.761 | 0.276 |
| 53 | Hypothyroidism NOS | Depression | 0.2626 | 3.75 | 0.075 |
| 54 | Hypothyroidism NOS | Anxiety disorder | 0.1782 | -0.015 | 0.078 |
| 55 | Hypothyroidism NOS | Tobacco use disorder | 0.1887 | -9.55 | -0.087 |

| | | | | | |
|---|---|---|---|---|---|
| 56 | Hypothyroidism NOS | Essential hypertension | 0.2626 | 8.593 | 0.234 |
| 57 | Hypothyroidism NOS | Myocardial infarction | 0.2582 | -3.644 | 0.028 |
| 58 | Hypothyroidism NOS | Coronary atherosclerosis | 0.2835 | -3.251 | 0.083 |
| 59 | Hypothyroidism NOS | Tachycardia NOS | 0.1313 | -8.516 | -0.105 |
| 60 | Hypothyroidism NOS | Asthma | 0.2196 | -0.56 | -0.014 |
| 61 | Hypothyroidism NOS | Diverticulosis | 0.1921 | -3.66 | 0.05 |
| 62 | Hypothyroidism NOS | Acute renal failure | 0.199 | -3.889 | -0.04 |
| 63 | Hypothyroidism NOS | Osteoarthrosis_localized_primary | 0.1484 | -3.436 | -0.003 |
| 64 | Myocardial infarction | Coronary atherosclerosis | 1.005 | 76.784 | 0.967 |
| 65 | Myocardial infarction | Tachycardia NOS | 0.2183 | 6.581 | 0.545 |
| 66 | Myocardial infarction | Asthma | 0.3295 | -4.192 | -0.174 |
| 67 | Myocardial infarction | Diverticulosis | 0.239 | -9.822 | -0.459 |
| 68 | Myocardial infarction | Acute renal failure | 0.6545 | 8.63 | 0.519 |
| 69 | Myocardial infarction | Osteoarthrosis_localized_primary | 0.197 | -11.483 | -0.4 |
| 70 | Obesity | Depression | 0.5422 | 7.959 | 0.173 |
| 71 | Obesity | Anxiety disorder | 0.3727 | 0.35 | 0.113 |
| 72 | Obesity | Tobacco use disorder | 0.6581 | 5.372 | 0.22 |
| 73 | Obesity | Essential hypertension | 0.5743 | 45.33 | 0.465 |
| 74 | Obesity | Myocardial infarction | 0.5183 | 3.859 | 0.23 |
| 75 | Obesity | Coronary atherosclerosis | 0.4578 | 6.488 | 0.278 |
| 76 | Obesity | Tachycardia NOS | 0.4843 | -7.794 | -0.044 |
| 77 | Obesity | Asthma | 0.4828 | 6.228 | -0.014 |
| 78 | Obesity | Diverticulosis | 0.3301 | -0.662 | 0.041 |
| 79 | Obesity | Acute renal failure | 0.903 | -1.981 | 0.037 |
| 80 | Obesity | Osteoarthrosis_localized_primary | 0.5754 | 8.665 | 0.047 |
| 81 | Tachycardia NOS | Asthma | 0.3411 | -8.608 | -0.278 |

| | | | | | |
|---|---|---|---|---|---|
| 82 | Tachycardia NOS | Diverticulosis | 0.2501 | -7.447 | -0.445 |
| 83 | Tachycardia NOS | Acute renal failure | 0.9687 | 11.372 | 0.703 |
| 84 | Tachycardia NOS | Osteoarthrosis_localized_primary | -0.06495 | -14.093 | -0.519 |
| 85 | Tobacco use disorder | Essential hypertension | 0.4827 | 17.496 | 0.223 |
| 86 | Tobacco use disorder | Myocardial infarction | 0.4979 | 21.49 | 0.292 |
| 87 | Tobacco use disorder | Coronary atherosclerosis | 0.4485 | 16.583 | 0.232 |
| 88 | Tobacco use disorder | Tachycardia NOS | 0.5459 | 4.613 | 0.386 |
| 89 | Tobacco use disorder | Asthma | 0.4426 | 0.5 | -0.073 |
| 90 | Tobacco use disorder | Diverticulosis | 0.3273 | -6.767 | -0.217 |
| 91 | Tobacco use disorder | Acute renal failure | 0.6529 | -1.649 | 0.193 |
| 92 | Tobacco use disorder | Osteoarthrosis_localized_primary | 0.3055 | -11.27 | -0.184 |
| 93 | Type 2 diabetes | Hyperlipidemia | 0.6138 | 43.785 | 0.527 |
| 94 | Type 2 diabetes | Obesity | 0.6616 | 40.865 | 0.497 |
| 95 | Type 2 diabetes | Depression | 0.3364 | -1.491 | -0.048 |
| 96 | Type 2 diabetes | Anxiety disorder | 0.2538 | -11.919 | -0.163 |
| 97 | Type 2 diabetes | Tobacco use disorder | 0.4777 | 0.799 | 0.122 |
| 98 | Type 2 diabetes | Essential hypertension | 0.5919 | 49.082 | 0.612 |
| 99 | Type 2 diabetes | Myocardial infarction | 0.4821 | 12.631 | 0.441 |
| 100 | Type 2 diabetes | Coronary atherosclerosis | 0.4775 | 22.387 | 0.469 |
| 101 | Type 2 diabetes | Tachycardia NOS | 0.5598 | -3.779 | 0.153 |
| 102 | Type 2 diabetes | Asthma | 0.3928 | -4.595 | -0.182 |
| 103 | Type 2 diabetes | Diverticulosis | 0.2513 | -8.522 | -0.191 |
| 104 | Type 2 diabetes | Acute renal failure | 0.6764 | 10.284 | 0.412 |
| 105 | Type 2 diabetes | Osteoarthrosis_localized_primary | 0.3886 | -8.424 | -0.168 |

### A15. Simulation study: Limited impact of phecode hierarchy on multimorbidity network conservation

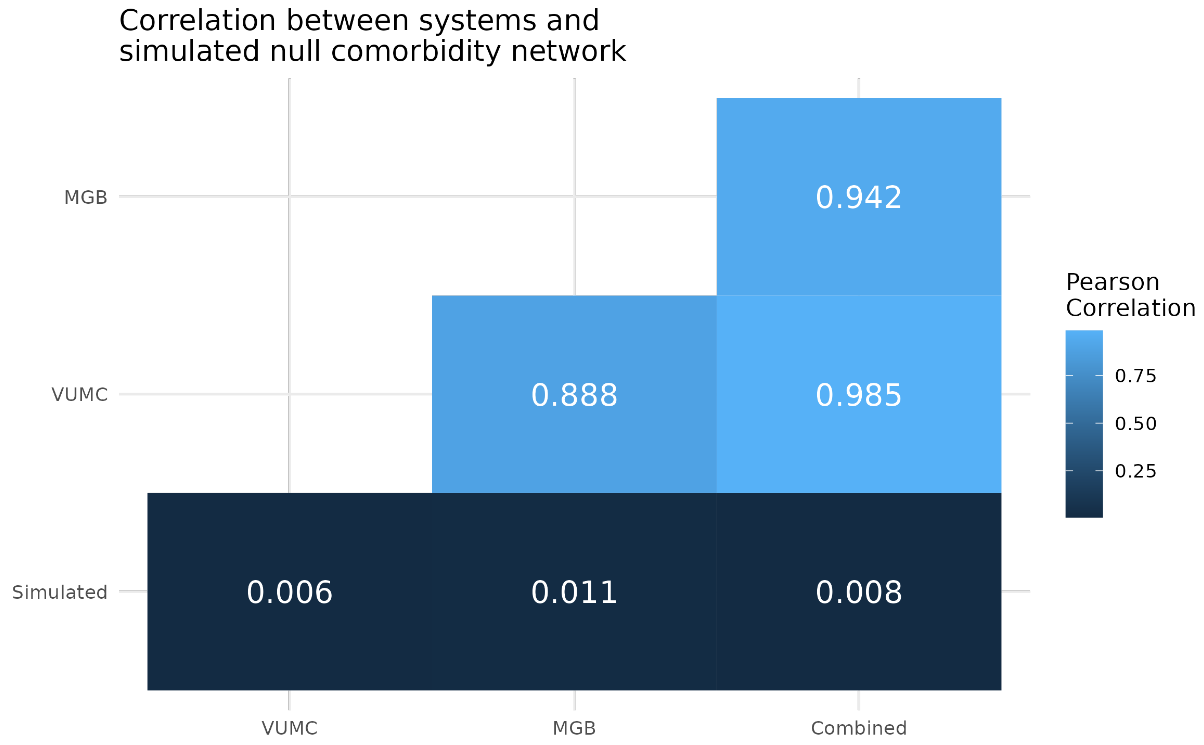

**Figure S12. Correlation heatmap: Multimorbidity conservation across VUMC, MGB, and simulated networks.** This heatmap compares the conservation of multimorbidity patterns between the VUMC, MGB, and a simulated network. The simulation, designed to isolate the effect of hierarchical phecode structure, used 5000 individuals with a 0.01 probability of exhibiting any given phecode within a defined neoplasm hierarchy. Logistic regression was used to model pairwise phecode relationships. The low correlation between the simulated network and both real-world networks suggests that the defined hierarchical structure alone has minimal influence on multimorbidity conservation patterns.

| Statistics for comorbidity z values with simulation | | | | | |
|---|---|---|---|---|---|
| | mean | sd | min | median | max |
| Vanderbilt | 5.142 | 10.389 | −9.162 | 1.287 | 127.653 |
| MGH | 3.454 | 9.720 | −12.220 | 0.266 | 92.816 |
| Simulated | 0.001 | 1.002 | −3.401 | −0.008 | 4.339 |

For speed of simulation, only codes in category *neoplasms* simulated

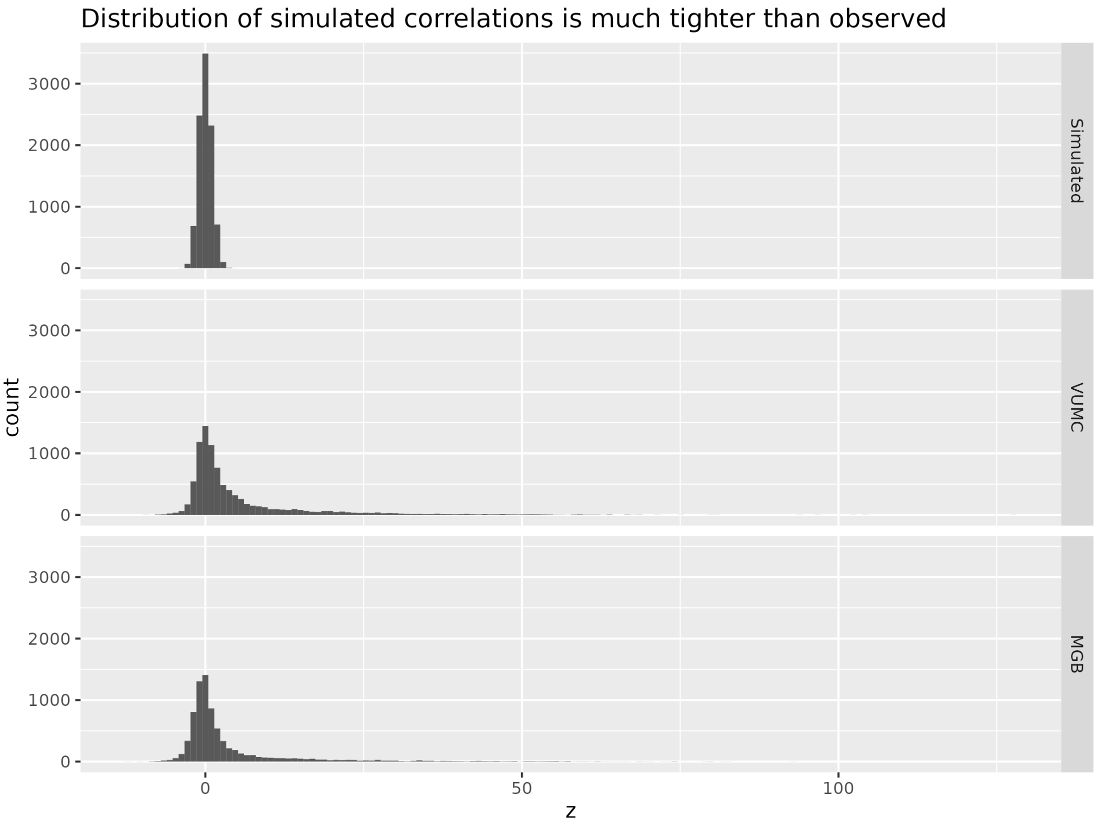

**Figure S13. Distribution of comorbidity strengths (z-values) in the simulated dataset.** The simulated dataset exhibits a narrower distribution of comorbidity strengths compared to the real-world datasets (VUMC and MGB), highlighting a key difference. VUMC or MGB.

#### A16. Top disease clusters

**Table S5. Cardiometabolic Cluster Phecodes from PheMIME**

| Phecode description | Phecode | Phecode category | degree |
|---|---|---|---|
| Mood disorders | 296.00 | mental disorders | 1,765 |
| Depression | 296.20 | mental disorders | 1,760 |
| Major depressive disorder | 296.22 | mental disorders | 1,700 |
| Substance addiction and disorders | 316.00 | mental disorders | 1,650 |
| Bipolar | 296.10 | mental disorders | 1,595 |
| Suicidal ideation or attempt | 297.00 | mental disorders | 1,587 |
| Alcoholic liver damage | 317.11 | mental disorders | 1,215 |
| Suicidal ideation | 297.10 | mental disorders | 1,337 |
| Schizophrenia and other psychotic disorders | 295.00 | mental disorders | 1,594 |
| Anxiety disorders | 300.00 | mental disorders | 1,765 |
| Alcohol-related disorders | 317.00 | mental disorders | 1,618 |
| Tobacco use disorder | 318.00 | mental disorders | 1,696 |
| Alcoholism | 317.10 | mental disorders | 1,587 |
| Generalized anxiety disorder | 300.11 | mental disorders | 1,582 |
| Posttraumatic stress disorder | 300.90 | mental disorders | 1,525 |
| Anxiety disorder | 300.10 | mental disorders | 1,753 |
| Cirrhosis of liver without mention of alcohol | 571.51 | digestive | 1,441 |
| Other chronic nonalcoholic liver disease | 571.50 | digestive | 1,644 |
| Esophageal bleeding (varices/hemorrhage) | 530.20 | digestive | 1,203 |
| Liver abscess and sequelae of chronic liver disease | 571.80 | digestive | 1,405 |
| Chronic liver disease and cirrhosis | 571.00 | digestive | 1,662 |

| | | | |
|---|---|---|---|
| Portal hypertension | 571.81 | digestive | 1,208 |
| Viral hepatitis | 070.00 | infectious diseases | 1,570 |
| Viral hepatitis C | 070.30 | infectious diseases | 1,522 |

**Table S6. Mental Health Cluster Phecodes from PheMIME**

| Phecode description | Phecode | Phecode category | degree |
|---|---|---|---|
| Back pain | 760.00 | symptoms | 1,771 |
| Cervicalgia | 761.00 | symptoms | 1,718 |
| Thoracic or lumbosacral neuritis or radiculitis, unspecified | 763.00 | symptoms | 1,614 |
| Cervical radiculitis | 765.00 | symptoms | 1,493 |
| Sciatica | 764.00 | symptoms | 1,582 |
| Dislocation | 830.00 | injuries & poisonings | 1,631 |
| Fracture of lower limb | 800.00 | injuries & poisonings | 1,653 |
| Fracture of tibia and fibula | 800.30 | injuries & poisonings | 1,424 |
| Fracture of vertebral column without mention of spinal cord injury | 805.00 | injuries & poisonings | 1,552 |
| Skull and face fracture and other intercranial injury | 819.00 | injuries & poisonings | 1,497 |
| Intracranial hemorrhage (injury) | 818.00 | injuries & poisonings | 1,367 |
| Fracture of upper limb | 803.00 | injuries & poisonings | 1,646 |
| Internal derangement of knee | 835.00 | injuries & poisonings | 1,630 |
| Fracture of ankle and foot | 801.00 | injuries & poisonings | 1,630 |
| Fracture of ribs | 807.00 | injuries & poisonings | 1,452 |
| Fracture of neck of femur | 800.10 | injuries & poisonings | 1,472 |
| Fracture of pelvis | 802.00 | injuries & poisonings | 1,308 |
| Fracture of clavicle or scapula | 803.30 | injuries & poisonings | 1,234 |
| Fracture of foot | 801.10 | injuries & poisonings | 1,500 |

| | | | |
|---|---|---|---|
| Fracture of radius and ulna | 803.20 | injuries & poisonings | 1,565 |
| Open wounds of head; neck; and trunk | 870.00 | injuries & poisonings | 1,651 |
| Subdural hemorrhage (injury) | 818.10 | injuries & poisonings | 1,200 |
| Fracture of hand or wrist | 804.00 | injuries & poisonings | 1,544 |
| Fracture of unspecified part of femur | 800.20 | injuries & poisonings | 1,309 |
| Fracture of unspecified bones | 809.00 | injuries & poisonings | 1,567 |
| Other open wound of head and face | 870.30 | injuries & poisonings | 1,428 |
| Sprains and strains | 840.00 | injuries & poisonings | 1,714 |
| Sprains and strains of back and neck | 841.00 | injuries & poisonings | 1,593 |
| Joint/ligament sprain | 840.30 | injuries & poisonings | 1,407 |
| Rotator cuff (capsule) sprain | 840.20 | injuries & poisonings | 1,417 |
| Spondylosis and allied disorders | 721.00 | musculoskeletal | 1,715 |
| Osteoarthrosis | 740.00 | musculoskeletal | 1,750 |
| Spondylosis without myelopathy | 721.10 | musculoskeletal | 1,708 |
| Intervertebral disc disorders | 722.00 | musculoskeletal | 1,732 |
| Peripheral enthesopathies and allied syndromes | 726.00 | musculoskeletal | 1,737 |
| Osteoarthritis; localized | 740.10 | musculoskeletal | 1,684 |
| Osteoarthrosis NOS | 740.90 | musculoskeletal | 1,745 |
| Pain in joint | 745.00 | musculoskeletal | 1,777 |
| Spinal stenosis | 720.00 | musculoskeletal | 1,671 |
| Displacement of intervertebral disc | 722.10 | musculoskeletal | 1,645 |
| Symptoms and disorders of the joints | 741.00 | musculoskeletal | 1,747 |
| Osteoarthrosis, localized, primary | 740.11 | musculoskeletal | 1,630 |
| Degeneration of intervertebral disc | 722.60 | musculoskeletal | 1,696 |
| Spinal stenosis of lumbar region | 720.10 | musculoskeletal | 1,616 |
| Other disorders of synovium, tendon, and bursa | 727.00 | musculoskeletal | 1,700 |
| Rupture of tendon, nontraumatic | 727.60 | musculoskeletal | 1,450 |
| Enthesopathy | 726.10 | musculoskeletal | 1,679 |
| Derangement of joint, non-traumatic | 742.00 | musculoskeletal | 1,528 |
| Other and unspecified disorders of back | 724.00 | musculoskeletal | 1,610 |
| Acquired spondylolisthesis | 738.40 | musculoskeletal | 1,364 |

| | | | |
|---|---|---|---|
| Other acquired musculoskeletal deformity | 738.00 | musculoskeletal | 1,510 |
| Intracranial hemorrhage | 430.00 | circulatory system | 1,556 |
| Subdural hemorrhage | 430.30 | circulatory system | 1,322 |
| Muscle/tendon sprain | 840.10 | injuries & poisonings | 1,162 |

**Table S7. Musculoskeletal Health Cluster Phecodes from PheMIME**

| Phecode description | Phecode | Phecode category | degree |
|---|---|---|---|
| Fever of unknown origin | 783.00 | symptoms | 1,771 |
| Disorders of lipoid metabolism | 272.00 | endocrine/metabolic | 1,779 |
| Hyperlipidemia | 272.10 | endocrine/metabolic | 1,777 |
| Type 1 diabetes | 250.10 | endocrine/metabolic | 1,617 |
| Type 2 diabetes with neurological manifestations | 250.24 | endocrine/metabolic | 1,566 |
| Diabetes mellitus | 250.00 | endocrine/metabolic | 1,744 |
| Type 2 diabetes | 250.20 | endocrine/metabolic | 1,741 |
| Insulin pump user | 250.30 | endocrine/metabolic | 1,585 |
| Polyneuropathy in diabetes | 250.60 | endocrine/metabolic | 1,445 |
| Encounter for long-term (current) use of anticoagulants | 286.20 | hematopoietic | 1,691 |
| Septal Deviations/Turbinate Hypertrophy | 470.00 | respiratory | 1,349 |
| Acute sinusitis | 464.00 | respiratory | 1,641 |
| Chronic sinusitis | 475.00 | respiratory | 1,673 |
| Acute upper respiratory infections of multiple or unspecified sites | 465.00 | respiratory | 1,765 |
| Acute bronchitis and bronchiolitis | 483.00 | respiratory | 1,670 |
| Shortness of breath | 512.70 | respiratory | 1,757 |
| Chronic pharyngitis and nasopharyngitis | 472.00 | respiratory | 1,566 |
| Allergic rhinitis | 476.00 | respiratory | 1,717 |
| Asthma | 495.00 | respiratory | 1,753 |
| Asthma with exacerbation | 495.20 | respiratory | 1,584 |
| Pneumonia | 480.00 | respiratory | 1,748 |

| | | | |
|---|---|---|---|
| Postnasal drip | 475.90 | respiratory | 947 |
| Acute pharyngitis | 465.20 | respiratory | 1,707 |
| Wheezing | 512.10 | respiratory | 1,564 |
| Cough | 512.80 | respiratory | 1,780 |
| Other dyspnea | 512.90 | respiratory | 1,752 |
| Other symptoms of respiratory system | 512.00 | respiratory | 1,789 |
| Other upper respiratory disease | 479.00 | respiratory | 1,590 |
| Eustachian tube disorders | 381.20 | sense organs | 1,269 |
| Otitis media and Eustachian tube disorders | 381.00 | sense organs | 1,648 |
| Otitis media | 381.10 | sense organs | 1,610 |
| Sensorineural hearing loss | 389.10 | sense organs | 1,498 |
| Suppurative and unspecified otitis media | 381.11 | sense organs | 1,564 |
| Other disorders of middle ear and mastoid | 385.00 | sense organs | 702 |
| Vertiginous syndromes and other disorders of vestibular system | 386.00 | sense organs | 1,451 |
| Impacted cerumen | 380.40 | sense organs | 1,599 |
| Cholesteatoma | 385.30 | sense organs | 600 |
| Dizziness and giddiness (Light-headedness and vertigo) | 386.90 | sense organs | 1,736 |
| Otalgia | 382.00 | sense organs | 1,571 |
| Hearing loss | 389.00 | sense organs | 1,665 |
| Conductive hearing loss | 389.20 | sense organs | 1,066 |
| Tinnitus | 389.40 | sense organs | 1,462 |
| Myocardial infarction | 411.20 | circulatory system | 1,671 |
| Cardiomyopathy | 425.00 | circulatory system | 1,625 |
| Rheumatic disease of the heart valves | 394.00 | circulatory system | 1,626 |
| Primary/intrinsic cardiomyopathies | 425.10 | circulatory system | 1,606 |
| Cardiac pacemaker in situ | 426.91 | circulatory system | 1,493 |
| Coronary atherosclerosis | 411.40 | circulatory system | 1,717 |
| Cardiac pacemaker/device in situ | 426.90 | circulatory system | 1,534 |
| Heart valve disorders | 395.00 | circulatory system | 1,731 |
| Ischemic Heart Disease | 411.00 | circulatory system | 1,741 |
| Cardiac defibrillator in situ | 426.92 | circulatory system | 1,337 |
| Cardiac conduction disorders | 426.00 | circulatory system | 1,698 |
| Nonspecific chest pain | 418.00 | circulatory system | 1,767 |
| Cardiomegaly | 416.00 | circulatory system | 1,684 |
| Nonrheumatic aortic valve disorders | 395.20 | circulatory system | 1,637 |
| Angina pectoris | 411.30 | circulatory system | 1,541 |
| Congestive heart failure (CHF) NOS | 428.10 | circulatory system | 1,703 |
| Unstable angina (intermediate coronary syndrome) | 411.10 | circulatory system | 1,477 |

| | | | |
|---|---|---|---|
| Atrioventricular [AV] block | 426.20 | circulatory system | 1,468 |
| Disease of tricuspid valve | 394.70 | circulatory system | 1,539 |
| Nonrheumatic mitral valve disorders | 395.10 | circulatory system | 1,686 |
| Abnormal heart sounds | 396.00 | circulatory system | 1,581 |
| Atrial fibrillation and flutter | 427.20 | circulatory system | 1,685 |
| Cardiac dysrhythmias | 427.00 | circulatory system | 1,775 |
| Atrial fibrillation | 427.21 | circulatory system | 1,682 |
| Other forms of chronic heart disease | 414.00 | circulatory system | 1,546 |
| Heart failure NOS | 428.20 | circulatory system | 1,548 |
| Hypertension | 401.00 | circulatory system | 1,780 |
| Congestive heart failure; nonhypertensive | 428.00 | circulatory system | 1,714 |
| Essential hypertension | 401.10 | circulatory system | 1,779 |
| Secondary/extrinsic cardiomyopathies | 425.20 | circulatory system | 1,436 |
| Paroxysmal tachycardia, unspecified | 427.10 | circulatory system | 1,610 |
| Other chronic ischemic heart disease, unspecified | 411.80 | circulatory system | 1,602 |
| Paroxysmal ventricular tachycardia | 427.12 | circulatory system | 1,484 |
| Heart valve replaced | 395.60 | circulatory system | 1,425 |
| Atrial flutter | 427.22 | circulatory system | 1,464 |
| Heart failure with reduced EF [Systolic or combined heart failure] | 428.30 | circulatory system | 1,461 |
| Sinoatrial node dysfunction (Bradycardia) | 427.80 | circulatory system | 1,465 |
| Cardiac congenital anomalies | 747.10 | congenital anomalies | 1,606 |
| Cardiac and circulatory congenital anomalies | 747.00 | congenital anomalies | 1,633 |
| Viral infection | 079.00 | infectious diseases | 1,685 |
| Valvular heart disease/ heart chambers | 747.12 | congenital anomalies | 1,253 |
| Other disorders of tympanic membrane | 384.00 | sense organs | 739 |
| Perforation of tympanic membrane | 384.40 | sense organs | 711 |
| Tympanosclerosis and middle ear disease related to otitis media | 385.50 | sense organs | 276 |

#### A17. EHR Multimorbidity network reveal clinical disease relationships, a case study on hydronephrosis

**Table S8. Most consistent conditions connected with hydronephrosis extracted from multimorbidity networks using PheMIME.**

| phecode | description | category | z_vandy | z_mgh | z_ukbb |
|---|---|---|---|---|---|
| 594.30 | Calculus of ureter | genitourinary | 61.092 | 54.604 | 65.698 |

| | | | | | |
|---|---|---|---|---|---|
| 594.00 | Urinary calculus | genitourinary | 60.345 | 54.618 | 10.429 |
| 586.40 | Stricture/obstruction of ureter | genitourinary | 59.798 | 53.876 | 78.531 |
| 594.10 | Calculus of kidney | genitourinary | 55.247 | 53.943 | 58.111 |
| 586.00 | Other disorders of the kidney and ureters | genitourinary | 56.279 | 49.681 | 32.203 |
| 751.30 | Obstructive genitourinary defect | congenital anomalies | 51.036 | 21.99 | 7.727 |
| 751.00 | Genitourinary congenital anomalies | congenital anomalies | 42.316 | 20.467 | |
| 590.00 | Pyelonephritis | genitourinary | 30.814 | 27.594 | 33.664 |
| 586.12 | Vesicoureteral reflux | genitourinary | 30.92 | 12.323 | 6.641 |
| 586.10 | Anatomical abnormatilies of kidney and ureters | genitourinary | 30.859 | 12.184 | |
| 189.20 | Cancer of bladder | neoplasms | 29.621 | 28.378 | 16.001 |
| 189.21 | Malignant neoplasm of bladder | neoplasms | 29.774 | 28.146 | 32.358 |
| 599.10 | Urinary obstruction | genitourinary | 31.475 | 23.343 | 20.776 |
| 591.00 | Urinary tract infection | genitourinary | 30.969 | 24.86 | 32.706 |
| 596.00 | Other disorders of bladder | genitourinary | 29.703 | 21.88 | 25.662 |
| 593.00 | Hematuria | genitourinary | 26.59 | 26.201 | 28.294 |
| 189.00 | Cancer of urinary organs (incl. kidney and bladder) | neoplasms | 26.093 | 25.567 | 23.57 |
| 751.20 | Congenital anomalies of urinary system | congenital anomalies | 27.415 | 12.119 | 6.127 |
| 594.80 | Renal colic | genitourinary | 25.7 | 21.176 | 32.788 |
| 597.20 | Urinary complications NEC | genitourinary | 24.125 | 22.156 | -0.341 |
| 599.00 | Other symptoms/disorders or the urinary system | genitourinary | 20.878 | 22.12 | 58.38 |
| 751.22 | Other specified congenital anomalies of kidney | congenital anomalies | 22.354 | 7.98 | 9.323 |
| 785.00 | Abdominal pain | symptoms | 22.01 | 16.841 | -0.427 |
| 198.00 | Secondary malignant neoplasm | neoplasms | 12.463 | 22.959 | 27.935 |
| 198.30 | Secondary malignant neoplasm of digestive systems | neoplasms | 12.243 | 21.837 | 33.25 |
| 596.50 | Functional disorders of bladder | genitourinary | 21.529 | 8.848 | 4.303 |

| | | | | | |
|---|---|---|---|---|---|
| 184.00 | Cancer of other female genital organs | neoplasms | 8.796 | 19.798 | 3.549 |
| 597.00 | Other disorders of urethra and urinary tract | genitourinary | 18.86 | 8.839 | 5.879 |
| 184.10 | Malignant neoplasm of ovary and other uterine adnexa | neoplasms | 7.856 | 18.675 | 9.884 |
| 599.20 | Retention of urine | genitourinary | 14.663 | 16.22 | 25.307 |
| 594.20 | Calculus of lower urinary tract | genitourinary | 17.293 | 12.521 | 20.277 |
| 560.00 | Intestinal obstruction without mention of hernia | digestive | 14.791 | 15.205 | |
| 585.00 | Renal failure | genitourinary | 15.19 | 14.688 | |
| 189.40 | Malignant neoplasm of other urinary organs | neoplasms | 15.642 | 13.206 | 21.576 |
| 180.10 | Cervical cancer | neoplasms | 11.985 | 16.051 | 11.674 |
| 184.11 | Malignant neoplasm of ovary | neoplasms | 6.646 | 16.666 | 26.524 |
| 182.00 | Malignant neoplasm of uterus | neoplasms | 7.687 | 16.472 | 14.184 |
| 593.10 | Gross hematuria | genitourinary | 15.202 | 11.895 | |
| 585.10 | Acute renal failure | genitourinary | 11.979 | 15.098 | 38.146 |
| 195.00 | Cancer_ suspected or other | neoplasms | 7.858 | 16.648 | 0.424 |
| 560.40 | Other intestinal obstruction | digestive | 11.013 | 15.358 | 16.905 |
| 185.00 | Cancer of prostate | neoplasms | 12.945 | 12.393 | 16.055 |
| 857.00 | Mechanical complication of unspecified genitourinary device_ implant_ and graft | injuries & poisonings | 8.655 | 15.184 | 38.83 |
| 041.40 | E. coli | infectious diseases | 13.195 | 9.459 | 15.905 |
| 751.21 | Cystic kidney disease | congenital anomalies | 13.763 | 6.923 | 4.604 |
| 579.00 | Other symptoms involving abdomen and pelvis | digestive | 11.219 | 13.654 | 18.149 |
| 597.10 | Urethral stricture (not specified as infectious) | genitourinary | 12.678 | 10.39 | 12.332 |
| 580.40 | Renal sclerosis_ NOS | genitourinary | 12.343 | 9.583 | |
| 592.12 | Chronic cystitis | genitourinary | 12.424 | 4.107 | 8.01 |
| 592.00 | Cystitis and urethritis | genitourinary | 12.299 | 6.585 | |
| 586.20 | Cyst of kidney_ acquired | genitourinary | 12.162 | 7.857 | 14.981 |
| 153.30 | Malignant neoplasm of rectum_ rectosigmoid junction_ and anus | neoplasms | 10.176 | 10.846 | 8.244 |

| | | | | | |
|---|---|---|---|---|---|
| 153.00 | Colorectal cancer | neoplasms | 9.43 | 11.234 | |
| 198.60 | Secondary malignancy of bone | neoplasms | 4.538 | 12.748 | 23.365 |
| 568.00 | Other disorders of peritoneum | digestive | 11.52 | 8.027 | 7.405 |
| 592.10 | Cystitis | genitourinary | 11.409 | 7.564 | 14.412 |
| 195.10 | Malignant neoplasm_ other | neoplasms | 6.088 | 12.837 | 12.077 |
| 189.10 | Cancer of kidney and renal pelvis | neoplasms | 10.886 | 9.51 | |
| 596.10 | Bladder neck obstruction | genitourinary | 8.773 | 11.44 | 6.833 |
| 198.40 | Secondary malignant neoplasm of liver | neoplasms | 7.17 | 11.691 | 17.195 |
| 567.00 | Peritonitis and retroperitoneal infections | digestive | 11.142 | 8.368 | 7.156 |
| 153.20 | Colon cancer | neoplasms | 9.014 | 10.257 | 6.575 |
| 198.10 | Secondary malignancy of lymph nodes | neoplasms | 7.36 | 11.125 | 21.722 |
| 585.30 | Chronic renal failure [CKD] | genitourinary | 12.514 | 6.509 | 15.454 |
| 560.10 | Paralytic ileus | digestive | 10.876 | 5.955 | 7.524 |
| 197.00 | Chemotherapy | neoplasms | 5.14 | 11.871 | |
| 568.10 | Peritoneal adhesions (postoperative) (postinfection) | digestive | 10.158 | 4.118 | 5.331 |
| 572.00 | Ascites (non malignant) | digestive | 9.694 | 7.979 | 9.521 |
| 990.00 | Effects radiation NOS | injuries & poisonings | 8.126 | 9.676 | 15.305 |
| 593.20 | Microscopic hematuria | genitourinary | 10.5 | 0.447 | |
| 559.00 | Ileostomy status | digestive | 8.747 | 8.612 | 12.08 |
| 585.20 | Renal failure NOS | genitourinary | 5.329 | 9.883 | 18.232 |
| 189.11 | Malignant neoplasm of kidney_ except pelvis | neoplasms | 8.42 | 7.76 | 5.339 |
| 580.00 | Nephritis; nephrosis; renal sclerosis | genitourinary | 10.109 | 4.205 | |
| 569.00 | Other disorders of intestine | digestive | 8.262 | 7.088 | 2.047 |
| 289.40 | Lymphadenitis | hematopoietic | 3.558 | 10.93 | 8.087 |
| 041.00 | Bacterial infection NOS | infectious diseases | 8.535 | 6.17 | 9.302 |
| 276.50 | Hypovolemia | endocrine/metabolic | 6.883 | 8.885 | 10.985 |
| 789.00 | Nausea and vomiting | symptoms | 6.245 | 9.445 | 5.705 |
| 589.00 | Abnormal results of function study of kidney | genitourinary | 7.096 | 7.842 | 6.965 |
| 599.40 | Urinary incontinence | genitourinary | 9.489 | 4.071 | -0.218 |
| 159.40 | Malignant neoplasm of retroperitoneum and peritoneum | neoplasms | 7.251 | 6.96 | 11.484 |
| 038.00 | Septicemia | infectious diseases | 6.178 | 8.025 | |
| 752.11 | Spina bifida | congenital anomalies | 7.837 | 2.786 | 2.654 |

| | | | | | |
|---|---|---|---|---|---|
| 159.00 | Malignant neoplasm of other and ill-defined sites within the digestive organs and peritoneum | neoplasms | 6.307 | 7.182 | 5.212 |
| 184.20 | Cancer of other female genital organs (excluding uterus and ovary) | neoplasms | 6.208 | 7.28 | 3.353 |
| 586.30 | Vascular disorders of kidney/hypertrophy | genitourinary | 7.444 | 5.126 | -0.708 |
| 587.00 | Kidney replaced by transpant | genitourinary | 6.768 | 6.198 | 6.385 |
| 599.30 | Dysuria | genitourinary | 7.497 | 4.195 | 4.074 |
| 994.20 | Sepsis | injuries & poisonings | 6.638 | 5.98 | 14.274 |
| 189.12 | Malignant neoplasm of renal pelvis | neoplasms | 6.01 | 6.738 | 5.839 |
| 752.10 | Neural tube defects | congenital anomalies | 6.875 | 3.207 | -1.231 |
| 180.00 | Cervical cancer and dysplasia | neoplasms | 6.554 | 5.918 | |
| 795.80 | Abnormal tumor markers | symptoms | 6.564 | -0.439 | |
| 614.32 | Chronic inflammatory pelvic disease | genitourinary | 6.463 | 5.398 | 3.916 |
| 599.80 | Other symptoms involving urinary system | genitourinary | 6.594 | 5.304 | 3.796 |
| 285.20 | Anemia of chronic disease | hematopoietic | 4.74 | 7.697 | 4.633 |
| 646.00 | Other complications of pregnancy NEC | pregnancy complications | 5.126 | 6.804 | -2.338 |
| 285.22 | Anemia in neoplastic disease | hematopoietic | 2.174 | 9.011 | 8.306 |
| 573.00 | Other disorders of liver | digestive | 5.592 | 6.29 | -0.259 |
| 994.00 | Sepsis and SIRS | injuries & poisonings | 5.877 | 5.982 | |
| 276.00 | Disorders of fluid_ electrolyte_ and acid-base balance | endocrine/metabolic | 5.502 | 6.463 | |
| 599.90 | Other abnormality of urination | genitourinary | 7.232 | 1.582 | |
| 038.10 | Gram negative septicemia | infectious diseases | 6.164 | 4.875 | -0.309 |
| 198.20 | Secondary malignancy of respiratory organs | neoplasms | 3.335 | 6.883 | 17.2 |
| 614.30 | Pelvic inflammatory disease (PID) | genitourinary | 6.749 | 2.799 | 1.834 |
| 452.00 | Other venous embolism and thrombosis | circulatory system | 4.345 | 6.713 | -0.325 |
| 853.00 | Complication of colostomy or enterostomy | injuries & poisonings | 6.598 | 3.765 | |
| 585.34 | Chronic Kidney Disease_ Stage IV | genitourinary | 7.139 | 3.554 | 8.363 |
| 187.00 | Cancer of other male genital organs | neoplasms | 2.865 | 6.981 | -0.699 |
| 401.22 | Hypertensive chronic kidney disease | circulatory system | 6.019 | 4.79 | 4.884 |
| 585.32 | End stage renal disease | genitourinary | 6.505 | 4.052 | 7.914 |
| 795.81 | Elevated carcinoembryonic antigen [CEA] | symptoms | 5.584 | -0.043 | |
| 569.20 | Gastrointestinal complications | digestive | 6.12 | 2.57 | 4.705 |

**Table S9. Most consistent conditions connected with hydronephrosis extracted from multimorbidity similarity networks using PheMIME**

| phecode | description | category | sim_vandy | sim_mgh | sim_ukbb |
|---|---|---|---|---|---|
| 594.30 | Calculus of ureter | genitourinary | 0.721 | 0.662 | 0.616 |
| 594.00 | Urinary calculus | genitourinary | 0.753 | 0.602 | 0.636 |
| 586.40 | Stricture/obstruction of ureter | genitourinary | 0.918 | 0.922 | 0.953 |
| 594.10 | Calculus of kidney | genitourinary | 0.771 | 0.67 | 0.657 |
| 586.00 | Other disorders of the kidney and ureters | genitourinary | 0.693 | 0.61 | 0.705 |
| 751.30 | Obstructive genitourinary defect | congenital anomalies | 0.803 | 0.7 | 0.455 |
| 751.00 | Genitourinary congenital anomalies | congenital anomalies | 0.652 | 0.567 | |
| 590.00 | Pyelonephritis | genitourinary | 0.738 | 0.723 | 0.749 |
| 586.12 | Vesicoureteral reflux | genitourinary | 0.647 | 0.505 | 0.502 |
| 586.10 | Anatomical abnormatilies of kidney and ureters | genitourinary | 0.633 | 0.473 | |
| 189.20 | Cancer of bladder | neoplasms | 0.676 | 0.669 | 0.504 |
| 189.21 | Malignant neoplasm of bladder | neoplasms | 0.677 | 0.665 | 0.517 |
| 591.00 | Urinary tract infection | genitourinary | 0.593 | 0.442 | 0.538 |
| 593.00 | Hematuria | genitourinary | 0.574 | 0.641 | 0.6 |
| 189.00 | Cancer of urinary organs (incl. kidney and bladder) | neoplasms | 0.653 | 0.706 | 0.537 |
| 594.80 | Renal colic | genitourinary | 0.598 | 0.603 | 0.539 |
| 198.00 | Secondary malignant neoplasm | neoplasms | 0.248 | 0.378 | 0.466 |
| 198.30 | Secondary malignant neoplasm of digestive systems | neoplasms | 0.355 | 0.472 | 0.447 |
| 184.00 | Cancer of other female genital organs | neoplasms | 0.366 | 0.507 | 0.323 |
| 184.10 | Malignant neoplasm of ovary and other uterine adnexa | neoplasms | 0.366 | 0.511 | 0.441 |
| 594.20 | Calculus of lower urinary tract | genitourinary | 0.692 | 0.636 | 0.713 |
| 560.00 | Intestinal obstruction without mention of hernia | digestive | 0.457 | 0.47 | |
| 585.00 | Renal failure | genitourinary | 0.323 | 0.226 | |
| 189.40 | Malignant neoplasm of other urinary organs | neoplasms | 0.536 | 0.544 | 0.645 |
| 180.10 | Cervical cancer | neoplasms | 0.439 | 0.564 | 0.477 |
| 184.11 | Malignant neoplasm of ovary | neoplasms | 0.334 | 0.493 | 0.458 |
| 182.00 | Malignant neoplasm of uterus | neoplasms | 0.348 | 0.493 | 0.381 |
| 593.10 | Gross hematuria | genitourinary | 0.708 | 0.662 | |
| 585.10 | Acute renal failure | genitourinary | 0.336 | 0.245 | 0.336 |
| 195.00 | Cancer_ suspected or other | neoplasms | 0.22 | 0.408 | 0.064 |
| 185.00 | Cancer of prostate | neoplasms | 0.529 | 0.519 | 0.433 |
| 857.00 | Mechanical complication of unspecified genitourinary device_ implant_ and graft | injuries & poisonings | 0.484 | 0.739 | 0.7 |

| | | | | | |
|---|---|---|---|---|---|
| 041.40 | E. coli | infectious diseases | 0.536 | 0.564 | 0.509 |
| 579.00 | Other symptoms involving abdomen and pelvis | digestive | 0.388 | 0.459 | 0.531 |
| 580.40 | Renal sclerosis_ NOS | genitourinary | 0.586 | 0.427 | |
| 592.12 | Chronic cystitis | genitourinary | 0.701 | 0.516 | 0.402 |
| 592.00 | Cystitis and urethritis | genitourinary | 0.571 | 0.439 | |
| 586.20 | Cyst of kidney_ acquired | genitourinary | 0.679 | 0.493 | 0.585 |
| 153.30 | Malignant neoplasm of rectum_ rectosigmoid junction_ and anus | neoplasms | 0.354 | 0.399 | 0.304 |
| 153.00 | Colorectal cancer | neoplasms | 0.358 | 0.431 | 0.222 |
| 198.60 | Secondary malignancy of bone | neoplasms | 0.187 | 0.311 | 0.389 |
| 568.00 | Other disorders of peritoneum | digestive | 0.449 | 0.454 | 0.46 |
| 592.10 | Cystitis | genitourinary | 0.601 | 0.499 | 0.49 |
| 195.10 | Malignant neoplasm_ other | neoplasms | 0.21 | 0.381 | 0.361 |
| 189.10 | Cancer of kidney and renal pelvis | neoplasms | 0.575 | 0.623 | |
| 198.40 | Secondary malignant neoplasm of liver | neoplasms | 0.264 | 0.398 | 0.396 |
| 567.00 | Peritonitis and retroperitoneal infections | digestive | 0.411 | 0.408 | 0.344 |
| 153.20 | Colon cancer | neoplasms | 0.359 | 0.432 | 0.288 |
| 198.10 | Secondary malignancy of lymph nodes | neoplasms | 0.212 | 0.346 | 0.371 |
| 585.30 | Chronic renal failure [CKD] | genitourinary | 0.285 | 0.164 | 0.179 |
| 560.10 | Paralytic ileus | digestive | 0.444 | 0.42 | 0.385 |
| 197.00 | Chemotherapy | neoplasms | 0.166 | 0.365 | |
| 568.10 | Peritoneal adhesions (postoperative) (postinfection) | digestive | 0.448 | 0.355 | 0.261 |
| 572.00 | Ascites (non malignant) | digestive | 0.355 | 0.375 | 0.289 |
| 593.20 | Microscopic hematuria | genitourinary | 0.539 | 0.301 | |
| 559.00 | Ileostomy status | digestive | 0.415 | 0.415 | 0.363 |
| 585.20 | Renal failure NOS | genitourinary | 0.325 | 0.256 | 0.378 |
| 189.11 | Malignant neoplasm of kidney_ except pelvis | neoplasms | 0.539 | 0.591 | 0.51 |
| 580.00 | Nephritis; nephrosis; renal sclerosis | genitourinary | 0.356 | 0.165 | |
| 569.00 | Other disorders of intestine | digestive | 0.331 | 0.354 | 0.177 |
| 289.40 | Lymphadenitis | hematopoietic | 0.152 | 0.313 | 0.275 |
| 041.00 | Bacterial infection NOS | infectious diseases | 0.292 | 0.248 | 0.367 |
| 276.50 | Hypovolemia | endocrine/metabolic | 0.341 | 0.43 | 0.372 |
| 589.00 | Abnormal results of function study of kidney | genitourinary | 0.411 | 0.335 | 0.357 |
| 159.40 | Malignant neoplasm of retroperitoneum and peritoneum | neoplasms | 0.331 | 0.468 | 0.445 |
| 038.00 | Septicemia | infectious diseases | 0.269 | 0.299 | -0.626 |

| | | | | | |
|---|---|---|---|---|---|
| 159.00 | Malignant neoplasm of other and ill-defined sites within the digestive organs and peritoneum | neoplasms | 0.323 | 0.404 | 0.261 |
| 184.20 | Cancer of other female genital organs (excluding uterus and ovary) | neoplasms | 0.322 | 0.366 | 0.212 |
| 586.30 | Vascular disorders of kidney/hypertrophy | genitourinary | 0.41 | 0.267 | 0.119 |
| 587.00 | Kidney replaced by transpant | genitourinary | 0.315 | 0.223 | 0.242 |
| 994.20 | Sepsis | injuries & poisonings | 0.285 | 0.266 | 0.396 |
| 189.12 | Malignant neoplasm of renal pelvis | neoplasms | 0.449 | 0.548 | 0.55 |
| 180.00 | Cervical cancer and dysplasia | neoplasms | 0.165 | 0.182 | |
| 795.80 | Abnormal tumor markers | symptoms | 0.336 | 0.367 | |
| 285.20 | Anemia of chronic disease | hematopoietic | 0.317 | 0.421 | 0.182 |
| 285.22 | Anemia in neoplastic disease | hematopoietic | 0.171 | 0.395 | 0.451 |
| 573.00 | Other disorders of liver | digestive | 0.343 | 0.358 | 0.115 |
| 994.00 | Sepsis and SIRS | injuries & poisonings | 0.26 | 0.274 | |
| 276.00 | Disorders of fluid_ electrolyte_ and acid-base balance | endocrine/metabolic | 0.291 | 0.308 | |
| 038.10 | Gram negative septicemia | infectious diseases | 0.371 | 0.347 | 0.149 |
| 198.20 | Secondary malignancy of respiratory organs | neoplasms | 0.203 | 0.304 | 0.396 |
| 452.00 | Other venous embolism and thrombosis | circulatory system | 0.228 | 0.233 | 0.212 |
| 585.34 | Chronic Kidney Disease_ Stage IV | genitourinary | 0.3 | 0.184 | 0.195 |
| 187.00 | Cancer of other male genital organs | neoplasms | 0.372 | 0.508 | 0.006 |
| 401.22 | Hypertensive chronic kidney disease | circulatory system | 0.269 | 0.187 | 0.156 |
| 585.32 | End stage renal disease | genitourinary | 0.281 | 0.148 | 0.218 |
| 795.81 | Elevated carcinoembryonic antigen [CEA] | symptoms | 0.321 | 0.264 | |
| 569.20 | Gastrointestinal complications | digestive | 0.294 | 0.255 | 0.343 |
| 285.00 | Other anemias | hematopoietic | 0.246 | 0.274 | 0.18 |
| 588.00 | Disorders resulting from impaired renal function | genitourinary | 0.323 | 0.189 | 0.145 |
| 783.00 | Fever of unknown origin | symptoms | 0.147 | 0.285 | 0.347 |
| 223.00 | Benign neoplasm of kidney and other urinary organs | neoplasms | 0.64 | 0.416 | 0.501 |
| 260.30 | Adult failure to thrive | endocrine/metabolic | 0.243 | 0.323 | |
| 038.30 | Bacteremia | infectious diseases | 0.23 | 0.289 | |
| 585.33 | Chronic Kidney Disease_ Stage III | genitourinary | 0.249 | 0.18 | 0.082 |
| 452.20 | Deep vein thrombosis [DVT] | circulatory system | 0.182 | 0.24 | |
| 585.40 | Chronic kidney disease_ Stage I or II | genitourinary | 0.299 | 0.164 | 0.064 |

| | | | | | |
|---|---|---|---|---|---|
| 187.20 | Malignant neoplasm of testis | neoplasms | 0.339 | 0.399 | 0.186 |
| 588.10 | Renal osteodystrophy | genitourinary | 0.316 | 0.253 | 0.106 |
| 614.31 | Acute inflammatory pelvic disease | genitourinary | 0.249 | 0.242 | 0.192 |
| 285.21 | Anemia in chronic kidney disease | hematopoietic | 0.306 | 0.166 | |
| 275.50 | Disorders of calcium/phosphorus metabolism | endocrine/metabolic | 0.264 | 0.212 | 0.362 |
| 260.00 | Protein-calorie malnutrition | endocrine/metabolic | 0.195 | 0.316 | 0.073 |
| 196.00 | Radiotherapy | neoplasms | 0.113 | 0.272 | |
| 276.13 | Hyperpotassemia | endocrine/metabolic | 0.319 | 0.161 | 0.33 |
| 750.22 | Congenital anomaly of gallbladder_ bile ducts_ liver_ pancreas | congenital anomalies | 0.264 | 0.211 | 0.2 |
| 560.30 | Peritoneal or intestinal adhesions | digestive | 0.365 | 0.418 | 0.304 |
| 260.20 | severe protein-calorie malnutrition | endocrine/metabolic | 0.257 | 0.191 | 0.055 |
| 592.11 | Acute cystitis | genitourinary | 0.508 | 0.213 | 0.461 |
| 041.20 | Streptococcus infection | infectious diseases | 0.315 | 0.272 | 0.273 |
| 750.00 | Digestive congenital anomalies | congenital anomalies | 0.202 | 0.152 | -0.007 |
| 980.00 | Encounter for long-term (current) use of antibiotics | infectious diseases | 0.201 | 0.262 | |
| 580.10 | Glomerulonephritis | genitourinary | 0.296 | 0.208 | |
| 080.00 | Postoperative infection | infectious diseases | 0.231 | 0.217 | 0.215 |
| 575.80 | Other disorders of biliary tract | digestive | 0.269 | 0.254 | 0.063 |
| 994.10 | Systemic inflammatory response syndrome (SIRS) | injuries & poisonings | 0.236 | 0.231 | 0.262 |
| 260.10 | Cachexia | endocrine/metabolic | 0.19 | 0.225 | 0.124 |
| 994.21 | Septic shock | injuries & poisonings | 0.23 | 0.227 | |
| 580.32 | Nephritis and nephropathy with pathological lesion | genitourinary | 0.307 | 0.175 | 0.145 |
| 585.31 | Renal dialysis | genitourinary | 0.25 | 0.128 | 0.211 |
| 550.60 | Incisional hernia | digestive | 0.357 | 0.19 | 0.24 |
| 170.00 | Cancer of bone and connective tissue | neoplasms | 0.151 | 0.294 | |
| 562.10 | Diverticulosis | digestive | 0.205 | 0.14 | 0.022 |
| 580.14 | Chronic glomerulonephritis_ NOS | genitourinary | 0.266 | 0.177 | 0.119 |
| 575.00 | Other biliary tract disease | digestive | 0.29 | 0.247 | |
| 580.13 | Acute glomerulonephritis_ NOS | genitourinary | 0.306 | 0.207 | 0.188 |
| 574.00 | Cholelithiasis and cholecystitis | digestive | 0.354 | 0.216 | |
| 275.00 | Disorders of mineral metabolism | endocrine/metabolic | 0.242 | 0.277 | 0.133 |

| | | | | | |
|---|---|---|---|---|---|
| 269.00 | Proteinuria | endocrine/metabolic | 0.264 | 0.119 | 0.182 |
| 586.11 | Small kidney | genitourinary | 0.327 | 0.163 | 0.555 |
| 562.00 | Diverticulosis and diverticulitis | digestive | 0.216 | 0.176 | |
| 187.10 | Malignant neoplasm of unspecified male genital organ | neoplasms | 0.336 | 0.443 | 0.23 |
| 220.00 | Benign neoplasm of ovary | neoplasms | 0.219 | 0.161 | 0.155 |
| 276.41 | Acidosis | endocrine/metabolic | 0.249 | 0.227 | 0.167 |
| 574.10 | Cholelithiasis | digestive | 0.334 | 0.176 | 0.041 |
| 157.00 | Pancreatic cancer | neoplasms | 0.282 | 0.325 | 0.212 |
| 170.20 | Cancer of connective tissue | neoplasms | 0.171 | 0.288 | 0.26 |
| 565.00 | Anal and rectal conditions | digestive | 0.184 | 0.223 | 0.092 |
| 851.00 | Complications of transplants and reattached limbs | injuries & poisonings | 0.222 | 0.174 | 0.186 |
| 588.20 | Secondary hyperparathyroidism (of renal origin) | genitourinary | 0.289 | 0.153 | |
| 560.20 | Impaction of intestine | digestive | 0.24 | 0.209 | 0.303 |
| 536.70 | Complications of gastrostomy_ colostomy and enterostomy | digestive | 0.171 | 0.267 | |
| 159.20 | Malignant neoplasm of small intestine_ including duodenum | neoplasms | 0.293 | 0.347 | 0.283 |
| 550.00 | Abdominal hernia | digestive | 0.326 | 0.199 | 0.236 |
| 151.00 | Cancer of stomach | neoplasms | 0.256 | 0.324 | 0.173 |
| 977.00 | Personal history of allergy to medicinal agents | injuries & poisonings | 0.178 | 0.259 | |
| 260.60 | Anorexia | endocrine/metabolic | 0.221 | 0.328 | 0.19 |
| 961.00 | Poisoning by other anti-infectives | injuries & poisonings | 0.209 | 0.268 | 0.064 |
| 514.20 | Solitary pulmonary nodule | respiratory | 0.22 | 0.197 | |
| 276.40 | Acid-base balance disorder | endocrine/metabolic | 0.23 | 0.194 | 0.069 |
| 288.20 | Elevated white blood cell count | hematopoietic | 0.207 | 0.187 | |
| 555.00 | Inflammatory bowel disease and other gastroenteritis and colitis | digestive | 0.208 | 0.255 | |
| 187.80 | Neoplasm of uncertain behavior of male genital organs | neoplasms | 0.207 | 0.345 | 0.209 |
| 275.53 | Disorders of phosphorus metabolism | endocrine/metabolic | 0.314 | 0.194 | 0.255 |
| 275.60 | Hypercalcemia | endocrine/metabolic | 0.231 | 0.219 | |
| 859.00 | Complication due to other implant and internal device | injuries & poisonings | 0.149 | 0.295 | 0.149 |
| 276.10 | Electrolyte imbalance | endocrine/metabolic | 0.235 | 0.201 | 0.166 |

| | | | | | |
|---|---|---|---|---|---|
| 288.11 | Neutropenia | hematopoietic | 0.124 | 0.222 | 0.29 |
| 550.40 | Umbilical hernia | digestive | 0.302 | 0.102 | 0.038 |
| 158.00 | Neoplasm of unspecified nature of digestive system | neoplasms | 0.27 | 0.291 | 0.199 |
| 211.00 | Benign neoplasm of other parts of digestive system | neoplasms | 0.212 | 0.113 | -0.136 |
| 112.00 | Candidiasis | infectious diseases | 0.178 | 0.248 | 0.214 |
| 963.00 | Poisoning by primarily systemic agents | injuries & poisonings | 0.129 | 0.291 | |
| 198.70 | Secondary malignant neoplasm of skin | neoplasms | 0.125 | 0.262 | 0.344 |
| 564.90 | Personal history of diseases of digestive system | digestive | 0.189 | 0.173 | -0.021 |
| 450.00 | Noninfectious disorders of lymphatic channels | circulatory system | 0.146 | 0.196 | 0.313 |
| 555.10 | Regional enteritis | digestive | 0.19 | 0.22 | 0.097 |
| 441.10 | Acute vascular insufficiency of intestine | circulatory system | 0.265 | 0.152 | 0.141 |
| 288.00 | Diseases of white blood cells | hematopoietic | 0.193 | 0.162 | 0.018 |
| 577.00 | Diseases of pancreas | digestive | 0.299 | 0.214 | 0.123 |
| 510.00 | Other diseases of lung | respiratory | 0.105 | 0.238 | 0.085 |
| 963.10 | Antineoplastic and immunosuppressive drugs causing adverse effects | injuries & poisonings | 0.136 | 0.285 | |
| 550.50 | Ventral hernia | digestive | 0.353 | 0.19 | 0.23 |
| 577.30 | Cyst and pseudocyst of pancreas | digestive | 0.284 | 0.13 | 0.075 |
| 288.10 | Decreased white blood cell count | hematopoietic | 0.138 | 0.222 | |
| 580.20 | Nephrotic syndrome without mention of glomerulonephritis | genitourinary | 0.208 | 0.093 | 0.271 |
| 289.30 | Personal history of diseases of blood and blood-forming organs | hematopoietic | 0.234 | 0.187 | |
| 194.00 | Cancer of other endocrine glands | neoplasms | 0.185 | 0.193 | 0.235 |
| 441.00 | Vascular insufficiency of intestine | circulatory system | 0.246 | 0.164 | 0.11 |
| 580.11 | Proliferative glomerulonephritis | genitourinary | 0.235 | 0.198 | 0.135 |
| 260.22 | Nutritional marasmus | endocrine/metabolic | 0.202 | 0.172 | 0.096 |
| 850.00 | Hemorrhage or hematoma complicating a procedure | injuries & poisonings | 0.194 | 0.122 | |
| 081.00 | Infection/inflammation of internal prosthetic device; implant; and graft | infectious diseases | 0.204 | 0.141 | 0.194 |
| 415.11 | Pulmonary embolism and infarction_ acute | circulatory system | 0.113 | 0.22 | 0.22 |
| 592.13 | Chronic interstitial cystitis | genitourinary | 0.183 | 0.299 | 0.32 |
| 271.90 | Other disorders of carbohydrate transport and metabolism | endocrine/metabolic | 0.203 | 0.099 | 0.108 |

| | | | | | |
|---|---|---|---|---|---|
| 008.52 | Intestinal infection due to C. difficile | infectious diseases | 0.21 | 0.232 | 0.372 |
| 575.20 | Obstruction of bile duct | digestive | 0.222 | 0.253 | 0.169 |
| 561.00 | Symptoms involving digestive system | digestive | 0.171 | 0.185 | -0.06 |
| 575.60 | Cholesterolosis of gallbladder | digestive | 0.27 | 0.074 | -0.007 |
| 008.50 | Bacterial enteritis | infectious diseases | 0.214 | 0.228 | -0.055 |
| 513.80 | Disorders of diaphragm | respiratory | 0.182 | 0.209 | 0.107 |
| 038.20 | Gram positive septicemia | infectious diseases | 0.19 | 0.106 | 0.109 |
| 255.00 | Disorders of adrenal glands | endocrine/metabolic | 0.194 | 0.135 | 0.127 |
| 283.21 | Hemolytic-uremic syndrome | hematopoietic | 0.182 | 0.192 | 0.151 |
| 275.30 | Disorders of magnesium metabolism | endocrine/metabolic | 0.164 | 0.31 | 0.239 |
| 555.20 | Ulcerative colitis | digestive | 0.177 | 0.218 | 0.064 |
| 270.38 | Other specified disorders of plasma protein metabolism | endocrine/metabolic | 0.286 | 0.183 | |
| 575.10 | Cholangitis | digestive | 0.223 | 0.236 | |
| 575.70 | Other disorders of gallbladder | digestive | 0.242 | 0.157 | 0.009 |
| 580.12 | Non-proliferative glomerulonephritis | genitourinary | 0.238 | 0.149 | 0.165 |
| 550.10 | Inguinal hernia | digestive | 0.277 | 0.136 | 0.042 |
| 571.00 | Chronic liver disease and cirrhosis | digestive | 0.224 | 0.075 | |
| 155.00 | Cancer of liver and intrahepatic bile duct | neoplasms | 0.219 | 0.294 | 0.177 |
| 514.00 | Abnormal findings examination of lungs | respiratory | 0.123 | 0.19 | 0.089 |
| 441.20 | Chronic vascular insufficiency of intestine | circulatory system | 0.213 | 0.038 | 0.026 |
| 170.10 | Bone cancer | neoplasms | 0.108 | 0.222 | 0.152 |
| 965.10 | Opiates and related narcotics causing adverse effects in therapeutic use | injuries & poisonings | 0.195 | 0.128 | |
| 458.20 | Iatrogenic hypotension | circulatory system | 0.171 | 0.143 | -0.164 |
| 537.00 | Other disorders of stomach and duodenum | digestive | 0.167 | 0.156 | -0.08 |
| 556.00 | Ulceration of the lower GI tract | digestive | 0.224 | 0.208 | 0.044 |
| 276.12 | Hyposmolality and/or hyponatremia | endocrine/metabolic | 0.215 | 0.091 | 0.128 |
| 276.14 | Hypopotassemia | endocrine/metabolic | 0.183 | 0.194 | 0.153 |
| 259.20 | Carcinoid syndrome | endocrine/metabolic | 0.219 | 0.284 | 0.294 |
| 155.10 | Malignant neoplasm of liver_ primary | neoplasms | 0.195 | 0.242 | 0.063 |
| 159.30 | Malignant neoplasm of gallbladder and extrahepatic bile ducts | neoplasms | 0.249 | 0.279 | 0.179 |
| 275.51 | Hypocalcemia | endocrine/metabolic | 0.161 | 0.276 | |

| | | | | | |
|---|---|---|---|---|---|
| 961.10 | Poisoning/allergy of sulfonamides | injuries & poisonings | 0.169 | 0.162 | 0.033 |
| 577.10 | Acute pancreatitis | digestive | 0.227 | 0.119 | 0.019 |
| 209.00 | Neuroendocrine tumors | neoplasms | 0.219 | 0.241 | |
| 573.50 | Jaundice (not of newborn) | digestive | 0.204 | 0.227 | 0.097 |
| 569.10 | Toxic gastroenteritis and colitis | digestive | 0.154 | 0.227 | 0.42 |
| 562.20 | Diverticulitis | digestive | 0.199 | 0.152 | |
| 316.10 | Polyneuropathy due to drugs | mental disorders | 0.155 | 0.277 | 0.372 |
| 592.20 | Urethritis and urethral syndrome | genitourinary | 0.215 | 0.132 | 0.386 |
| 289.50 | Diseases of spleen | hematopoietic | 0.223 | 0.264 | 0.167 |
| 555.21 | Ulcerative colitis (chronic) | digestive | 0.177 | 0.189 | 0.048 |
| 556.10 | Ulceration of intestine | digestive | 0.204 | 0.179 | 0.046 |
| 540.00 | Appendiceal conditions | digestive | 0.219 | 0.261 | 0.14 |
| 283.20 | Non-autoimmune hemolytic anemias | hematopoietic | 0.151 | 0.183 | 0.192 |
| 281.90 | Deficiency anemias | hematopoietic | 0.193 | 0.063 | -0.012 |
| 540.11 | Acute appendicitis | digestive | 0.175 | 0.209 | 0.138 |
| 540.10 | Appendicitis | digestive | 0.181 | 0.24 | 0.093 |
| 150.00 | Cancer of esophagus | neoplasms | 0.164 | 0.165 | 0.12 |

#### A18. PheMIME: Interactive web application to explore phenome-wide multimorbidities

We have provided an interactive web-application to explore multimorbidity patterns across multiple EHR systems along with the phenotype’s relative position within the broader comorbidity network (https://prod.tbilab.org/PheMIME/) (Zhang et al., 2023). This application provides pairwise morbidity patten information with comparisons across institutions, it also shows the disease’s relative position within the combined multimorbidity networks, examines the reproducibility compared across multiple EHR systems, and explores the subgraph structure of the comorbidity networks using associationSubgraphs (Strayer et al., 2022). We provide demonstration using Hydronephrosis (595.00) as an example.
